# Spatio-temporal dynamics of snakebite envenoming in Mato Grosso do Sul, Brazil (2007–2023): environmental interfaces, occupational risk, and predictive modeling

**DOI:** 10.1101/2025.11.11.25339998

**Authors:** Mitzy Stephanny Machado, André Valério da Silva, Juliano Yasuo Oda, Aline Rafaela da Silva Rodrigues Machado, Alex Martins Machado

## Abstract

Snakebite envenoming remains a significant public health concern in Brazil. This study analyzed the epidemiological, spatial, and environmental patterns of snakebite envenoming in Mato Grosso do Sul (MS) between 2007 and 2023. An average of 415 cases were reported annually, showing no consistent long-term trend but a clear seasonal peak between January and April. Most incidents were caused by *Bothrops* species, which were widely distributed across the state, whereas *Crotalus*, *Micrurus*, and *Lachesis* occurred less frequently and exhibited more geographically restricted distributions. Spatial analyses revealed pronounced heterogeneity among municipalities, with persistent high-risk areas in central-southern and northwestern regions, including Campo Grande, Aquidauana, and Bonito. Demographically, most cases occurred in adult men of mixed ethnicity, though children and elderly individuals showed greater vulnerability to severe outcomes. Lower limbs were the most frequently affected anatomical sites, and rapid access to antivenom therapy contributed to low overall mortality. Environmental and land-use variables, particularly agriculture, pastures, and urban areas, were positively associated with snakebite envenoming, whereas biome and climate exerted limited influence. Municipalities with high tourism activity also exhibited elevated case numbers, reflecting interactions between human mobility, exposure, and reporting. These findings indicate that snakebite risk in MS is primarily shaped by local socio-environmental factors, with strong seasonal and spatial variability, and provide evidence-based guidance for targeted prevention, equitable resource allocation, and strategic public health planning focused on vulnerable populations. These findings can inform predictive surveillance and guide regional antivenom distribution strategies

## Introduction

The implementation of the United Nations’ 2030 Agenda and the development of the Sustainable Development Goals (SDGs) established a global commitment to transform living conditions on the planet, both in the present and for future generations. This agenda highlights and integrates different social, environmental, and economic dimensions, recognizing that progress in one area can drive improvements in several others. Among the major obstacles to achieving these goals are the neglected tropical diseases, which disproportionately affect vulnerable populations in low- and middle-income countries. Among these diseases, snakebite envenoming stands out due to its significant humanitarian burden. Although it is not an infectious disease, it constitutes a serious health condition resulting from venomous snakebites **[1,2]**.

According to World Health Organization (WHO) estimates, between 4.5 and 5.4 million snakebite cases occur globally each year, resulting in 1.8 to 2.7 million cases of clinical envenoming and 81,000 to 138,000 deaths annually, in addition to around 400,000 cases of permanent disabilities **[3]**. More recent studies, however, suggest substantial underreporting, indicating up to 5.5 million cases per year and more than 120,000 annual deaths **[4]**. The Global Burden of Disease analysis estimated a mortality of over 63,000 cases (2019 CI: 38,900–78,600), reflecting variability among data sources but reaffirming the likelihood of significant underreporting **[5]**. Regional disparities are particularly pronounced in sub-Saharan Africa and Southeast Asia. A study conducted within the Association of Southeast Asian Nations (ASEAN) reported an annual average of 242,648 cases (95% CI: 209,810–291,023), with over 15,000 deaths (95% CI: 7,592– 33,949) and 954 amputations (95% CI: 383–1,797). Similarly, an analysis across 41 sub-Saharan African countries identified 268,471 (95% CI: 221,868–314,782) annual cases, with 12,290 (95% CI: 9,710– 14,924) deaths and 14,766 (95% CI: 9,043–25,183) amputations **[6,7]**.

Although the greatest global burden of snakebite envenoming is concentrated in Africa and South Asia, the Americas also experience a substantial and heterogeneous impact. Several Latin American countries, such as Brazil, Venezuela, Ecuador, Bolivia, Paraguay, French Guiana, and Guyana, and Caribbean nations including Panama and Nicaragua report high incidence rates in rural areas and among vulnerable populations. Despite significant mortality, underreporting remains a major challenge, hindering direct comparison between countries. Nonetheless, historical series and local studies clearly indicate that snakebite envenoming continues to cause thousands of deaths and hundreds of thousands of permanent disabilities across the region **[4,5].**

Brazil reports one of the highest absolute numbers of snakebite envenoming in the Americas, with systematic notifications to the Ministry of Health through the Information System for Notifiable Diseases (Sistema de Informação de Agravos de Notificação – SINAN) and the provision of antivenom through the Unified Health System (Sistema Único de Saúde – SUS). Although these measures have improved case management, local inequalities in healthcare access persist, increasing the risk of severe cases and long-term sequelae. Geospatial and epidemiological studies show that sociodemographic factors, healthcare service coverage, variations in species distribution (e.g., *Bothrops*, *Crotalus*, *Lachesis*, *Micrurus*), as well as environmental characteristics, account for much of the heterogeneity observed within the country **[8,9]**.

Pará and Amazonas, located in the northern region of Brazil, account for the highest number of reported snakebite cases in the country (80,581 and 16,936 cases between 2007 and 2023, respectively). Although much of the research has focused on factors such as climate, deforestation, social vulnerability, and exposed population groups, a significant gap remains in studies conducted in other regions of Brazil **[10–13]**. According to recent surveys, the Central-West region presents an intermediate incidence of cases compared with the North and Northeast regions; however, this does not imply lower epidemiological relevance **[14,15]**. In the state of Mato Grosso do Sul (MS), located in the Central-West region, specifically, recent years have seen an expansion of agricultural and livestock activities, increased deforestation, and the intensification of ecotourism. These factors, combined with climatic variations and the coexistence of three major biomes (Pantanal, Cerrado, and Atlantic Forest), have expanded the human–snake interface, thereby increasing the risk of snakebite cases **[16]**.

The landscape of snakebite envenoming in MS is characterized by spatial heterogeneity and a strong connection to local environmental and economic conditions. Distinct municipal patterns are observed within the state, with variation associated with municipality size and the distribution of existing biomes (Pantanal, Cerrado, and remnants of the Atlantic Forest). These patterns reinforce that rural areas and human–nature interface zones present higher risks. Regional analyses in MS demonstrate that, although the average incidence is lower than in parts of the Amazon, local risk may be elevated in municipalities with greater vegetation loss, agricultural activity, and habitat fragmentation **[17,18]**.

The incorporation of quantitative spatiotemporal modeling methods has proven fundamental to understanding the dynamic patterns of neglected diseases, including snakebite envenoming, particularly in regions strongly influenced by environmental and seasonal factors **[15]**. Integrated autoregressive moving average (ARIMA) models and their seasonal extensions (SARIMA) have been widely used in epidemiological surveillance to estimate trends, forecast incidence fluctuations, and identify potential high-risk periods **[19,20]**. These approaches allow the integration of temporal dependence and, in the case of SARIMA, capture annual cycles and climatic patterns that may modulate snakebite occurrence, such as variations in rainfall, temperature, and agricultural activity dynamics.

In addition, spatial analysis combined with count regression models, such as the negative binomial model, makes it possible to assess the contribution of environmental and socioeconomic factors, thereby strengthening predictive capacity and surveillance planning. In this context, the integrated application of these techniques in MS enables not only the description of historical patterns of snakebite cases but also the projection of short-term risk scenarios, contributing to more effective and spatially sensitive public health policies.

Although previous studies have characterized the national epidemiological profile of snakebite envenoming, research focusing specifically on MS remains scarce, particularly studies that integrate environmental, sociodemographic, occupational, and temporal determinants over long-term periods. This gap limits the ability to develop targeted public health interventions and anticipate seasonal or spatial risk patterns. Expanding such investigations is crucial to understanding how local environmental conditions, land-use patterns, and human activities interact to influence snakebite cases. Accordingly, this study aims to describe the spatial and temporal distribution of snakebite cases in MS between 2007 and 2023, evaluating associations with land use, climate, biome, and tourism classification. Furthermore, we apply robust count models to quantify these relationships and project short-term temporal trends, providing evidence to support more effective, spatially sensitive planning, surveillance, and prevention strategies.

## Materials and Methods

### Data sources

The data used in this study were obtained from various public and official databases, integrated at the municipal level for the state of MS, Brazil (357,125 km² – 2.9 million inhabitants). The unit of analysis was the municipality, totaling 79 administrative units that comprise the state (Figure 1). Given its extensive territory and environmental diversity, MS provides an ideal context for assessing spatial and temporal patterns of snakebite envenoming.

**Figure 1.**
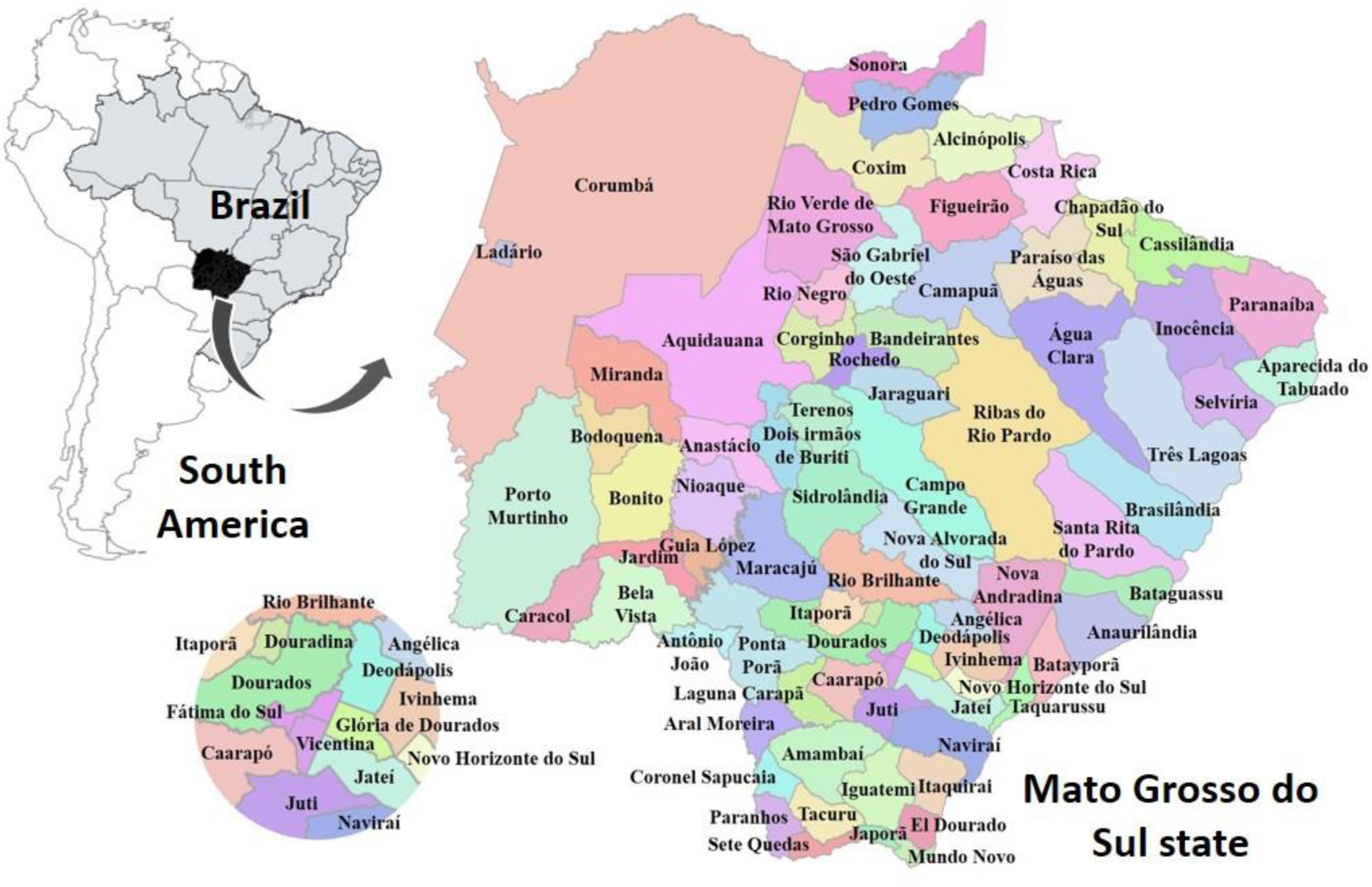
Geographical location of the study area. The map shows Brazil’s position within South America, highlighting the state of Mato Grosso do Sul. On the right, an enlarged view of the state displays its municipal boundaries and the names of the 79 municipalities that compose the analyzed territory. This representation provides spatial context for the areas mentioned throughout the text, facilitating geographical identification for readers who may not be familiar with the region.

### Ethical considerations

The data used in this study were derived from the Information System for Notifiable Diseases (Sistema de Informação de Agravos de Notificação – SINAN/DATASUS), which are publicly accessible secondary data, anonymized and without individual identification.

Therefore, submission to a Research Ethics Committee was not required, in accordance with Resolution CNS No. 510/2016, which waives ethical review for studies using public-domain information. It is noteworthy that although part of the analysis included the self-reported variable “race/color,” which contains the category “Indigenous,” all data were treated exclusively in aggregated form and without any linkage to specific communities, ensuring complete confidentiality and respect for Indigenous populations.

### Snakebite cases

Records of snakebite cases caused by venomous snakes (*Bothrops*, *Crotalus*, *Lachesis*, and *Micrurus*) were obtained from SINAN **[21]**, available through the DATASUS platform (Ministry of Health, Brazil). Data collection was carried out between May and July 2025, including all reported cases from January 2007 to December 2023.

The selected variables included municipality of occurrence, year, snake genus, sex, age group, race/color, anatomical site of the bite, time to medical care, antivenom administration, outcome, and work-relatedness (occupational accident). Data were extracted, consolidated, and processed using R software, with duplicate records and inconsistent entries in key variables removed.

### Biomes and climate

The delimitation of predominant biomes (Cerrado, Pantanal, and Atlantic Forest) was obtained from the official Brazilian Biomes shapefile provided by the Brazilian Institute of Geography and Statistics (IBGE) [22]. Climatic classifications were derived from the Köppen–Geiger Climate Classification, as adapted by the IBGE [23], which defines three main climatic types in the state: Hot-humid, Sub-hot-humid, and Semi-hot-humid. Complementary climatic variables (mean annual temperature and accumulated precipitation) were retrieved from the National Institute of Meteorology (INMET) database for the period 2007–2023. For analytical purposes, each municipality was assigned to its predominant biome and climate type according to the spatial predominance criterion, in which the class covering more than 50% of the municipal area determined its classification. This approach ensured consistency between administrative boundaries and environmental reference units.

### Tourism classification

Tourism classification data were obtained from the Mapa do Turismo Brasileiro (Brazilian Tourism Map) published by the Ministry of Tourism **[24]**. Municipalities were grouped into categories A to E (or “uncategorized”) according to their economic performance and visitor flow. The most recent version of this classification was used, and categories were linked to municipalities according to official codes.

### Land use and land cover

Land use and cover data were extracted from the MapBiomas Collection 8 dataset **[25]**, which provides area estimates (km²) of different land-cover classes for each municipality. The following classes were considered: artificial areas, agricultural areas, pasture, forest mosaic, planted forest (silviculture), forest vegetation, wetland, grassland, grassland mosaic, and continental water. These areas were consolidated at the municipal level and, when necessary, adjusted proportionally to the total municipal area for relative comparisons.

### Indigenous land

Indigenous territories were mapped using official data from the National Foundation of Indigenous Peoples (Fundação Nacional dos Povos Indígenas -FUNAI) **[26]**. These polygons were spatially overlaid with municipal boundaries to identify intersections, without any individual or community-level identifiers. The data were used exclusively to assess geographic overlap with high-incidence areas, ensuring complete confidentiality and respect for Indigenous populations.

### Spatial integration and processing

All spatial datasets were harmonized under the SIRGAS 2000 / UTM Zone 21S reference system using QGIS 3.36.1. Geoprocessing operations, such as re-projection, vector overlay, and area calculation, were performed within this environment. Tabular data were integrated using the official municipal code to ensure consistency across sources. Final maps were produced in QGIS using standardized color schemes and symbols to represent case counts and the genus of the snake responsible for each incident.

### Statistical analysis

Statistical analyses were conducted in R software (version 4.5.1) **[27]**. Initially, descriptive statistics (means, standard deviations, and frequencies), graphical representations, and spatial maps were produced to explore the distribution of snakebite cases across climatic, biome, tourism, and land-use categories. Differences between groups were tested using non-parametric Kruskal–Wallis tests, followed, when significant, by post-hoc Dunn tests with Bonferroni correction for multiple comparisons. Differences in proportions (e.g., occupational vs. non-occupational cases) were evaluated using proportion tests.

To assess temporal trends and produce short-term forecasts, autoregressive integrated moving average (ARIMA) models were fitted to annual time series of reported cases **[19,28]**. Automatic parameter selection (p, d, q) was performed using R’s built-in functions based on the Akaike Information Criterion (AIC). Series stationarity was assessed using the Augmented Dickey–Fuller (ADF) test, and differencing was applied when required. Models were estimated for the entire state (2007–2023) and for selected municipalities with informative time series. Forecasts were generated for 2024–2026 with 80% and 95% confidence intervals. Model diagnostics included residual inspection, autocorrelation function (ACF) analysis, and the Ljung–Box test to verify the absence of residual autocorrelation.

Seasonal patterns were further analyzed using monthly aggregated data (2007–2023). The series was structured with monthly frequency (ts, frequency = 12) and subjected to graphical inspection and STL decomposition to identify trend, seasonal, and irregular components. Stationarity was evaluated using the ADF test. Seasonal autoregressive integrated moving average (SARIMA) models were fitted by maximum likelihood using the forecast package in R (stepwise = FALSE; approximation = FALSE) **[19,28]**. Model selection was based on the lowest corrected AIC (AICc), parameter significance, and diagnostic checks using residual ACF and Ljung–Box tests. Forecasts were generated for 24 months (2024–2025) with 80% and 95% confidence intervals, yielding projections of expected monthly incidence in the post-observation period. Predictive performance was assessed using RMSE, MAE, and MAPE.

Associations between snakebite cases and continuous land-use variables were initially examined using Spearman’s correlation. Simple and multiple linear regression models (using both absolute area in km² and relative percentages) were then fitted to evaluate relationships between case numbers and land-use classes. To jointly assess the effects of categorical variables (tourism, climate, and biome) and continuous land-use variables on the total number of cases, Poisson regression models were fitted. Overdispersion was tested by the ratio of Pearson residuals to degrees of freedom; when detected, negative binomial models were applied as a robust alternative. Incidence Rate Ratios (IRR) and corresponding 95% confidence intervals were calculated for model interpretation.

Prior to model fitting, multicollinearity among explanatory variables was assessed using the Variance Inflation Factor (VIF), and no variable exceeded the threshold value of 5, indicating acceptable independence for inclusion in the models. Integrative models combining different variable sets (e.g., tourism and land use) were compared with the global model using the Akaike Information Criterion (AIC) to evaluate model fit and parsimony. Model diagnostics included residual analysis, evaluation of influential observations using Cook’s distance, and inspection of dispersion parameters to confirm model adequacy and the absence of influential outliers.

## Results

Between 2007 and 2023, a total of 7,063 cases of snakebite envenoming were reported in the state of MS, corresponding to an average of approximately 415 cases per year. The distribution of reports was not homogeneous over time. Although the first year analyzed (2007) recorded only 267 notifications, the number of cases increased substantially in the following two years (2008 and 2009), reaching an average of more than 500 annual cases. A decrease in notifications was observed during the COVID-19 pandemic period (2020–2022), with 340, 335, and 302 cases, respectively, and this downward trend persisted in 2023, with only 296 reported cases (**Figure 2A**).

**Figure 2.**
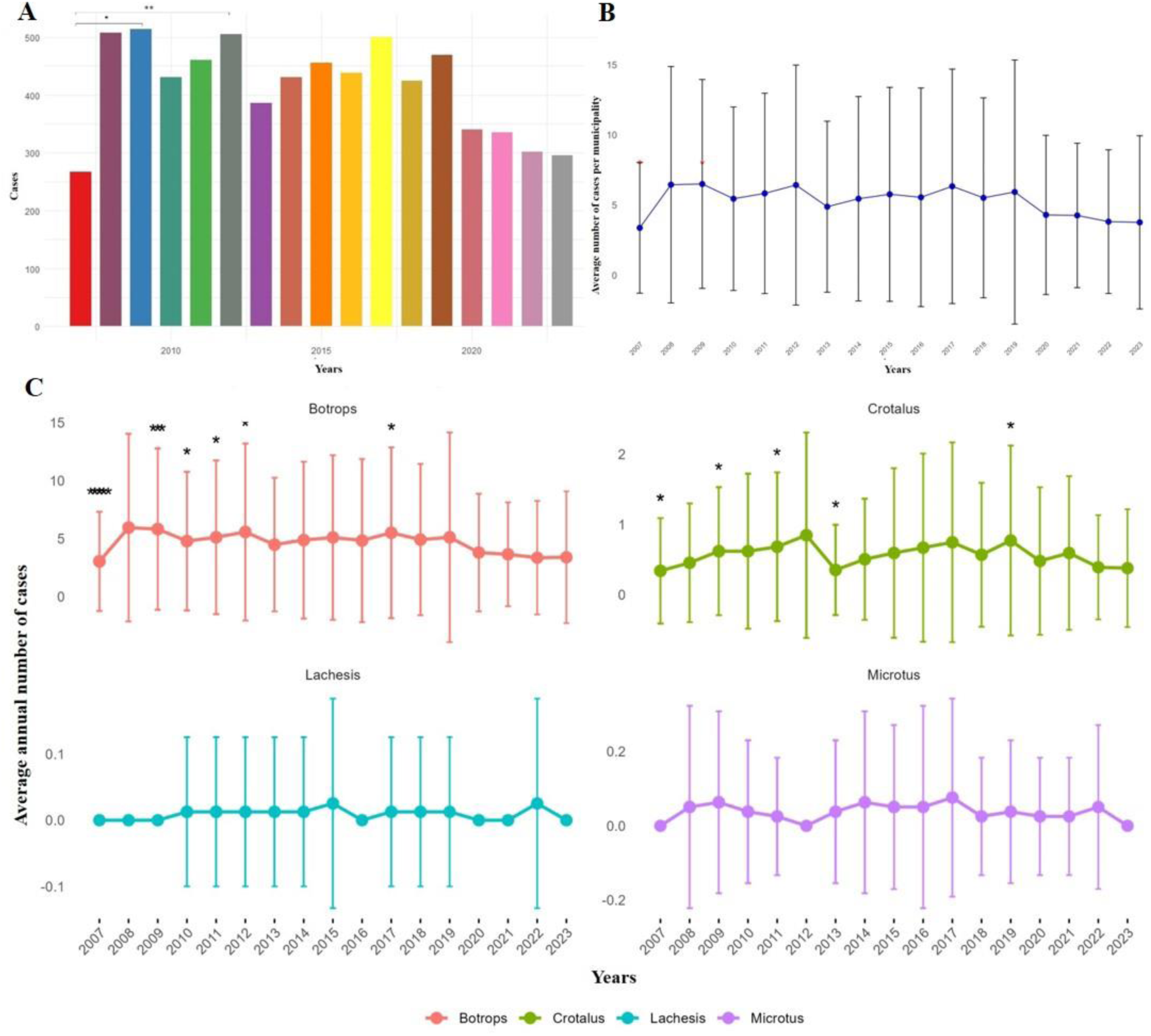
Temporal distribution of reported snakebite incidents in Mato Grosso do Sul (2007–2023). (A) Annual variation in the total number of cases, showing an initial increase until 2009 and a reduction during the pandemic period (2020–2023). Significant differences were observed among the evaluated years (Kruskal–Wallis, χ² = 31.99; df = 16; p = 0.01), particularly between 2007 and 2009 (Dunn’s test, p = 0.023). (B) Temporal trend of cases, indicating a significant but low-magnitude decline over the study period (coefficient = –0.082; p = 0.037; adjusted R² = 0.0025). (C) Annual means and standard deviations by snake genus, showing predominance of *Bothrops*, followed by *Crotalus*, *Micrurus*, and *Lachesis*. Significant differences were found only for *Bothrops* (χ² = 30.87; df = 16; p = 0.0139), and regression analysis indicated a non-significant declining trend (β = –6.50; p = 0.0619), suggesting overall stability among genera.

A year-by-year comparison of snakebite occurrences between 2007 and 2023 indicated statistically significant differences among years (Kruskal–Wallis, χ² = 31.99; df = 16; p = 0.01). Post-hoc Dunn tests with Bonferroni correction for multiple comparisons revealed a significant difference only between 2007 and 2009 (p = 0.023), while no other pairwise comparisons were statistically significant. These findings suggest that, although annual fluctuations occurred, most years did not differ significantly from one another. A simple linear regression trend analysis indicated a small but statistically significant decrease in the number of cases over time (coefficient = –0.082; p = 0.037). However, the model’s explanatory power was low (adjusted R² = 0.0025), indicating that temporal variation alone does not adequately explain case variability (**Figure 2B**).

The univariate time series analysis using an AutoRegressive Integrated Moving Average (ARIMA) model showed that the ARIMA(0,1,0) configuration best fit the data, indicating the absence of a consistent directional trend and characterizing the series as a random walk, in which annual variations reflect stochastic fluctuations around the historical mean. The model demonstrated good fit (AIC = 189.3; MAPE = 13.0%) and residuals without significant autocorrelation (Ljung–Box test, p = 0.719), suggesting model adequacy. Accordingly, the annual number of cases remained relatively stable between 2007 and 2023, with no significant increasing or decreasing trend. Forecasts for 2024–2026 suggest the continuation of current levels (approximately 300 cases per year), with wide confidence intervals (95% CI: 133–463 cases in 2024), reflecting high interannual variability and the absence of a clear temporal pattern or consistent trend.

The monthly time series analysis revealed a well-defined seasonal pattern in snakebite occurrence in MS. The augmented Dickey–Fuller (ADF) test indicated stationarity of the series (p < 0.01). The selected model, SARIMA (3,0,0)(2,1,0)(12), showed a good fit to the observed data (RMSE = 8.92; MAPE = 23.4%), although a slight residual autocorrelation was detected according to the Ljung–Box test (p = 0.0066). The series decomposition indicated that the highest number of cases consistently occurred between January and April, whereas the lowest values were concentrated between June and August, reflecting the influence of regional climatic seasonality on snake activity and human exposure in rural environments.

Complementarily, **Figure 2C** presents the annual means and standard deviations of snakebite cases by genus for the period 2007–2023. *Bothrops* bites were the most frequent throughout the entire period, with apparent interannual fluctuations (means ranging from 3.03 to 5.94 cases per year per municipality). The genus *Crotalus* ranked second in frequency, although with substantially lower annual means (0.34–0.84 cases/year/municipality) and interannual variability. *Micrurus* ranked third, with no recorded cases in 2007, 2012, and 2023, and very low annual means (0.00–0.07 cases/year/municipality). Finally, *Lachesis* presented the lowest values, with no cases recorded in 2007–2009, 2016, 2020, and 2021, and mean values between 0.00 and 0.02 cases/year/municipality. These variations were assessed using the Kruskal–Wallis test, which indicated statistically significant differences only for *Bothrops* (p < 0.05).

The temporal analysis of *Bothrops* bites revealed heterogeneity among years (Kruskal–Wallis χ² = 30.87; df = 16; p = 0.0139). The post-hoc Dunn test indicated that 2007 exhibited significantly lower values compared to several subsequent years (e.g., 2009, 2011, 2012, 2017), although most differences lost significance after Bonferroni correction. A simple linear regression suggested a slight decreasing trend over the study period (β = –6.50 cases/year; p = 0.0619), though not statistically significant. For the genera *Crotalus* (χ² = 20.25; p = 0.2094), *Micrurus* (χ² = 19.82; p = 0.2281), and *Lachesis* (χ² = 10.79; p = 0.8221), no statistically significant differences were detected among years. Despite minor annual fluctuations, these variations did not represent consistent temporal trends, likely reflecting the low frequency of such accidents in the state and the consequently limited statistical power to detect subtle differences.

In addition to temporal variations among snake genera, the spatial analysis revealed relevant differences between municipalities in the state. These disparities were first assessed through statistical comparisons between municipalities, later summarized in risk maps, and finally explored in detail through geoprocessing, which integrates the temporal and spatial distribution of cases by species. The comparative analysis among municipalities revealed highly significant differences in the occurrence of snakebite cases (Kruskal–Wallis, χ² = 850.78; df = 78; p < 0.001). The post hoc Dunn test with Bonferroni adjustment revealed marked contrasts between municipalities with high and low incidence rates, highlighting the pronounced spatial heterogeneity of snakebite occurrence across the state. Significant differences were observed primarily between high-incidence municipalities in the central-southern region (e.g., Campos Grande and Aquidauana) and those with lower records in the eastern portion (e.g., Anaurilândia and Batayporã). Conversely, some municipalities displayed similar incidence patterns, suggesting local homogeneity in certain areas. These findings underscore that, while snakebite distribution is markedly uneven, groups of municipalities share comparable levels of occurrence. Spatial mapping further clarifies these disparities and supports the identification of regional risk patterns.

Geoprocessing analyses integrating temporal and spatial dimensions confirmed heterogeneous distribution patterns over time. Areas with persistently high numbers of cases were concentrated in the central-southern (Campo Grande, Dourados, and Rio Brilhante) and northwestern regions (Corumbá and Coxim), whereas the eastern region exhibited sporadic or time-restricted occurrences (**Figures 3–5**). Cluster analysis identified three distinct profiles: Cluster 1 comprised municipalities with the highest incidence (Campo Grande and Aquidauana); Cluster 2 included the majority with low to moderate occurrence (e.g., Anastácio and Bonito), and Cluster 3 represented mid-sized regional centers with intermediate to high incidence levels (e.g., Dourados, Corumbá and Sidrolândia). Together, these clusters reinforce the intra-state heterogeneity of snakebite distribution (**Figure S1**).

**Figure 3.**
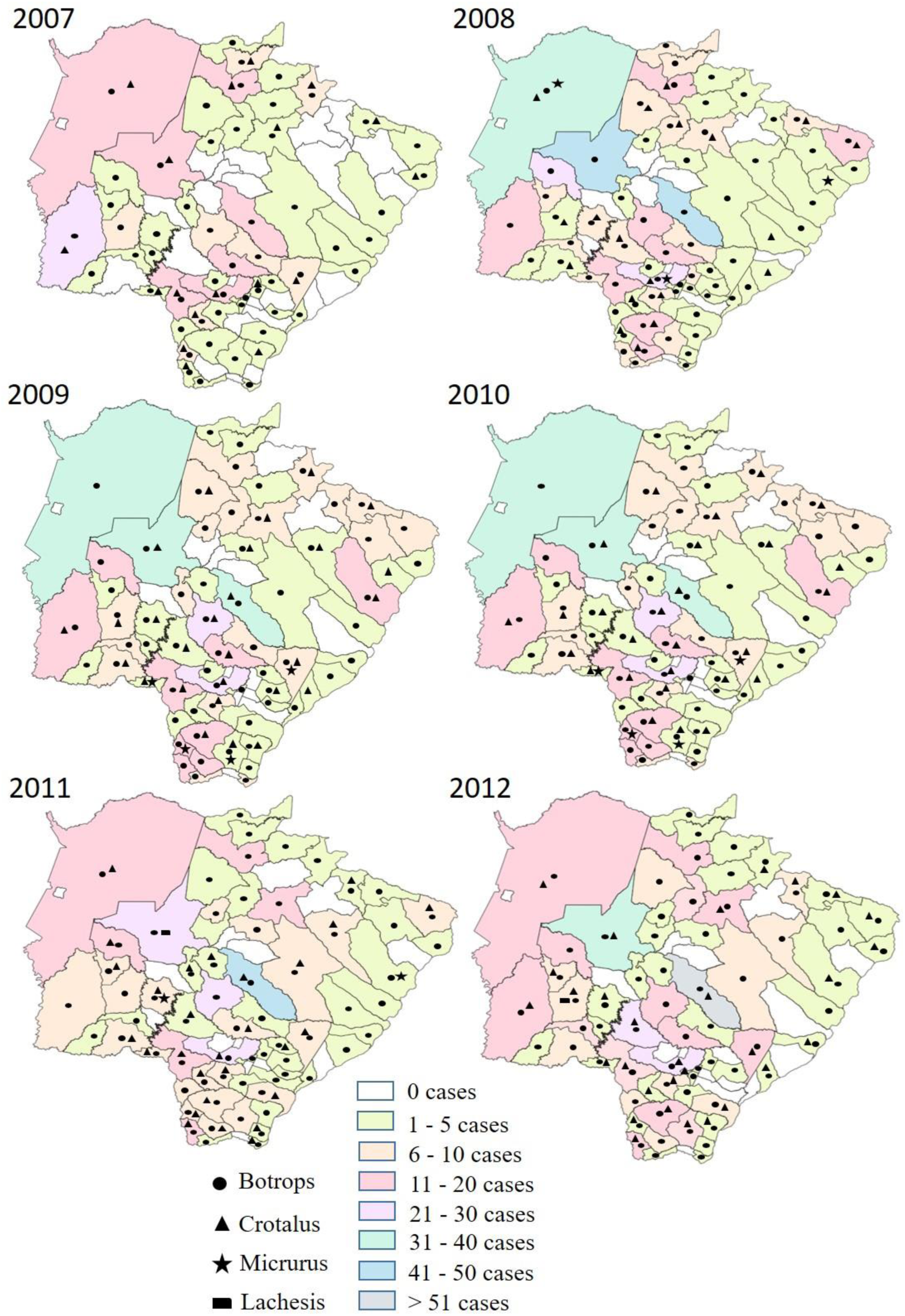
Spatiotemporal distribution of snakebite envenoming by snake genus in the municipalities of Mato Grosso do Sul, 2007–2012. The map depicts geoprocessed annual records per municipality, highlighting areas with persistent high incidence during the initial years of the time series, particularly in the south-central and northwestern regions of the state.

**Figure 4.**
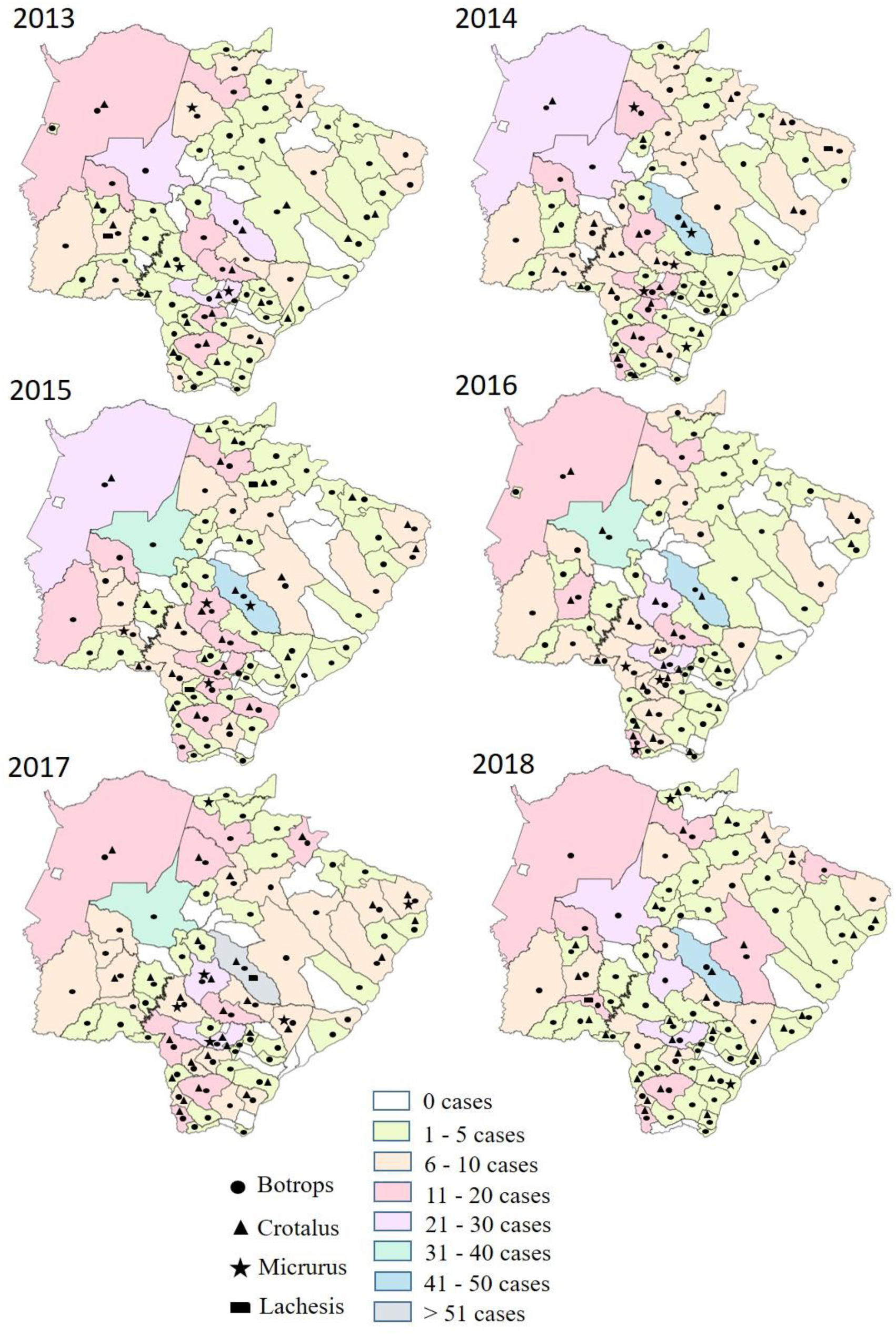
Spatiotemporal distribution of snakebite envenomings by snake genus in the municipalities of Mato Grosso do Sul, 2013–2018. Persistence of high-incidence foci is observed in the south-central and northwestern regions, with localized expansion of intermediate-occurrence areas, reflecting the continued epidemiological vulnerability of these regions.

**Figure 5.**
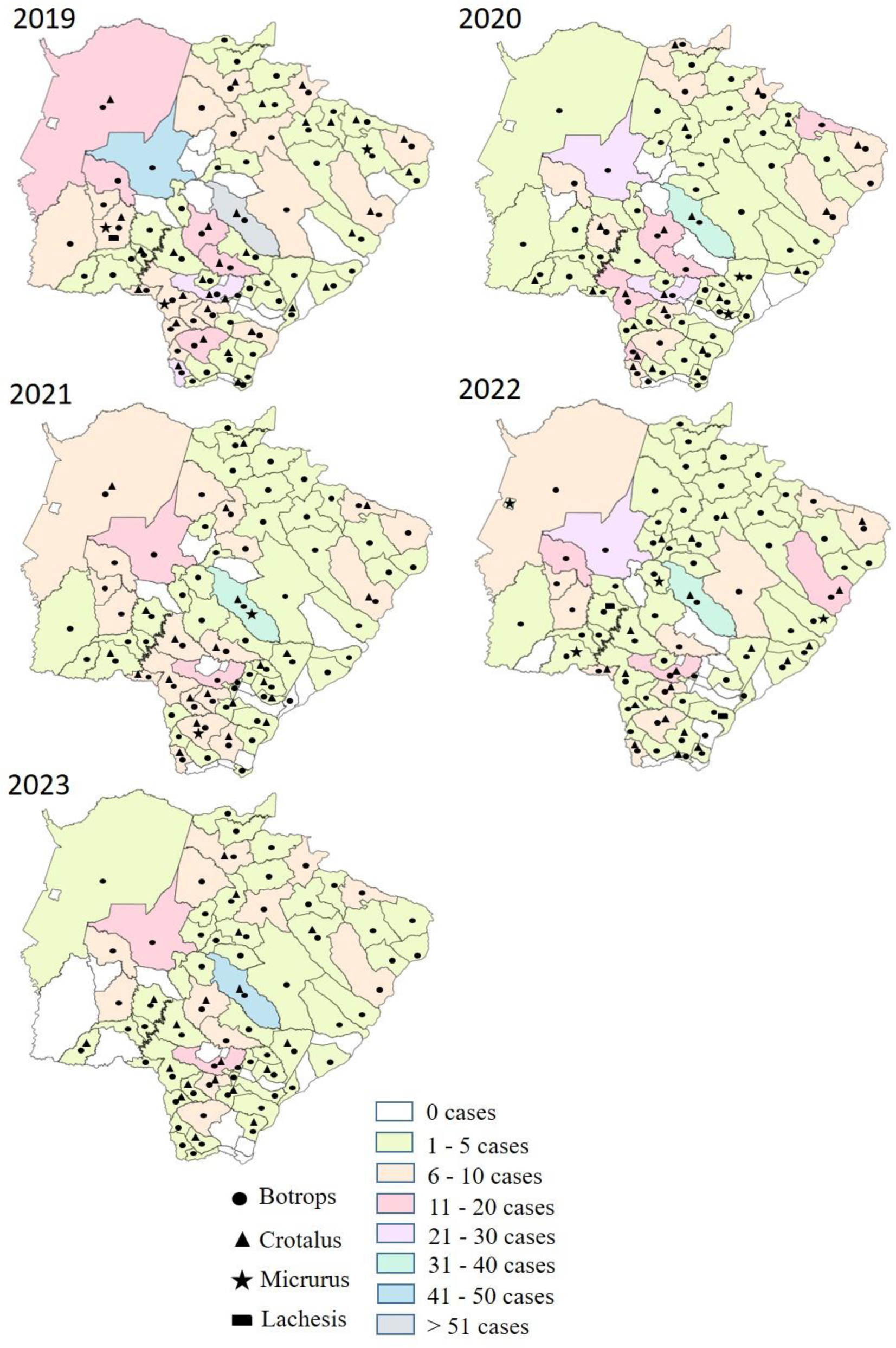
Spatiotemporal distribution of snakebite envenomings by snake genus in the municipalities of Mato Grosso do Sul, 2019–2023. High numbers of cases persisted in Campo Grande, Dourados, Aquidauana, and neighboring municipalities, despite an overall decline in total cases. This pattern suggests spatial stability of high-risk foci, with interannual variation likely influenced by the pandemic period and local environmental factors.

The *Bothrops* genus showed the widest distribution, accounting for the majority of snakebite envenoming and occurring in nearly all municipalities with reported incidents, except Fátima do Sul and Japorã, where the total number of records was very low. This genus was more frequent in medium- and large-sized municipalities, representing the main contributor to the overall snakebite burden in the state. The *Crotalus* genus, although also widely distributed, was absent in some municipalities, such as Jateí, Rochedo, and Anastácio, indicating a less homogeneous spatial pattern. In contrast, *Micrurus* and *Lachesis* displayed a restricted spatial distribution and low frequency. *Micrurus* cases were primarily concentrated in the central-southern region (e.g., Campo Grande, Dourados, Sidrolândia, and Nova Andradina) and, to a lesser extent, in the northwestern region (Corumbá and Rio Verde de Mato Grosso), with sporadic records elsewhere. *Lachesis* was reported in only nine municipalities, with a notable concentration in Bonito, an important tourist destination in the southwestern region, where, despite the small absolute number of cases, the relative risk was comparatively high in specific years (**Figures 3–5**). Overall, the spatial distribution of snakebites was highly heterogeneous across municipalities and genera, underscoring the diversity of epidemiological settings within the state.

Cluster analysis based on genus composition identified four profiles. In all clusters, *Bothrops* predominated, with average counts ranging from low (≈47 cases) to very high (≈464). *Crotalus* played a secondary role, with higher values observed in clusters with elevated overall incidence (up to 57.5 cases), whereas *Lachesis* contributed minimally, restricted to a few municipalities.

To summarize these discrepancies visually, a risk map was developed to classify municipalities according to the relative intensity of snakebite cases by genus (**Figure 6**). These maps illustrate the relative risk of each genus compared with the total number of cases within each municipality. Municipalities such as Campo Grande and Aquidauana exhibited the highest relative risk for *Bothrops* bites, whereas *Crotalus*, despite having fewer total cases, showed a broader range of municipalities with high-risk categories, including Campo Grande, Dourados, and Antônio João. *Micrurus* also presented elevated risk areas in the central-southern region, particularly in Dourados, Caarapó, Sidrolândia, and Nova Andradina. Interestingly, *Lachesis* exhibited an overall low risk consistent with its low frequency but stood out in Bonito, where the scarcity of other genera made its few records relatively more prominent.

**Figure 6.**
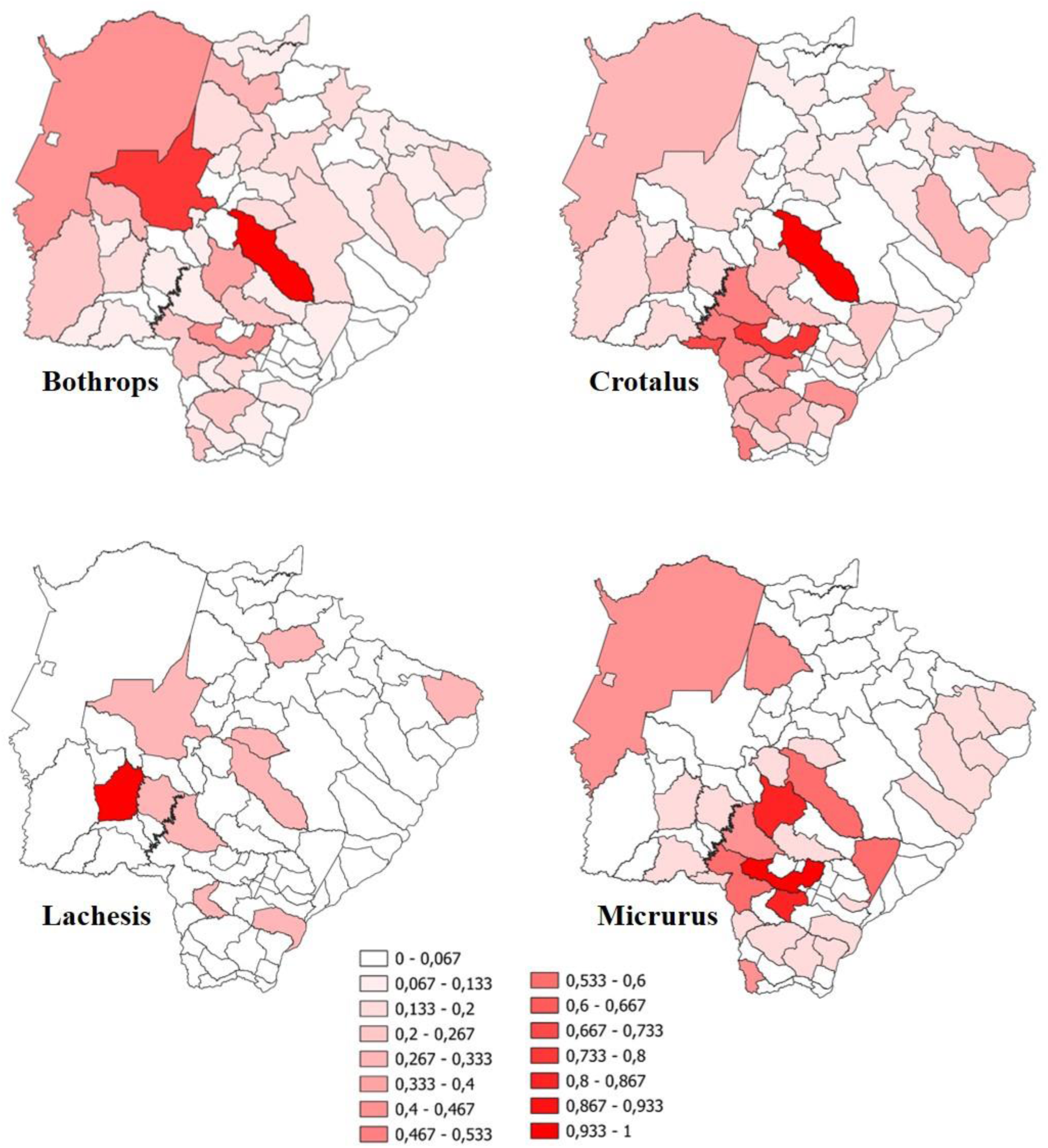
Spatial distribution and relative risk of snakebite envenomings by snake genus in the municipalities of Mato Grosso do Sul (2007–2023). The heat map classifies municipalities according to the relative intensity of cases by genus (proportion of accidents by each genus relative to the municipal total), revealing marked spatial heterogeneity. *Bothrops* accounted for the highest risks, mainly concentrated in Campo Grande and Aquidauana. *Crotalus* cases were prominent in Campo Grande, Dourados, and Antônio João, while *Micrurus* showed high-risk areas in the south-central region, particularly Dourados, Caarapó, Sidrolândia, and Nova Andradina. *Lachesis* displayed low relative risk, with a localized focus in Bonito. The observed heterogeneity reflects distinct ecological and occupational conditions across state regions.

The forecasts obtained from the ARIMA models indicate overall stability in the expected number of snakebite cases for the next three years (2024–2026) in municipalities with the highest case counts. In Campo Grande, the model estimated a mean of 41.5 annual cases (95% CI: 19.3–63.6), with no significant indication of either an increasing or decreasing trend. In Aquidauana, the predicted mean was 28.9 annual cases (95% CI: 10.4–47.5), also suggesting stability. Similar patterns were observed in Dourados and Corumbá, with variations limited to the uncertainty bounds. Sidrolândia, on the other hand, showed a slight upward tendency, possibly associated with recent agricultural expansion and population growth (**Figure S2A**). Overall, the results indicate maintenance of the levels observed in the historical series, with no strong evidence of short-term growth or decline. However, monthly projections (SARIMA model) for 2024–2025 confirmed the persistence of the seasonal pattern, with predicted peaks of approximately 36–42 cases during the first four months of the year and declines to 12–15 cases during the dry season (**Figure S2B**).

Regarding sex, a clear predominance of male cases was observed: 5,408 in men (76.5%) compared with 1,664 in women (23.5%), corresponding to a ratio of approximately 3:1 (p < 0.001). Temporal analysis showed that this difference remained consistent over time, reflecting the occupational and exposure-related profile commonly reported for snakebite cases in other regions of Brazil (Figure 7A). When analyzed by race, most cases occurred among *pardo* individuals, as classified by the Brazilian census (42.6%), followed by white (29.4%) and Indigenous (16.3%) individuals. This pattern may reflect the spatial overlap between Indigenous lands (**Figure S3**) and areas of higher snakebite incidence, as well as barriers to timely medical care, and therefore warrants particular attention in public health policies.

**Figure 7.**
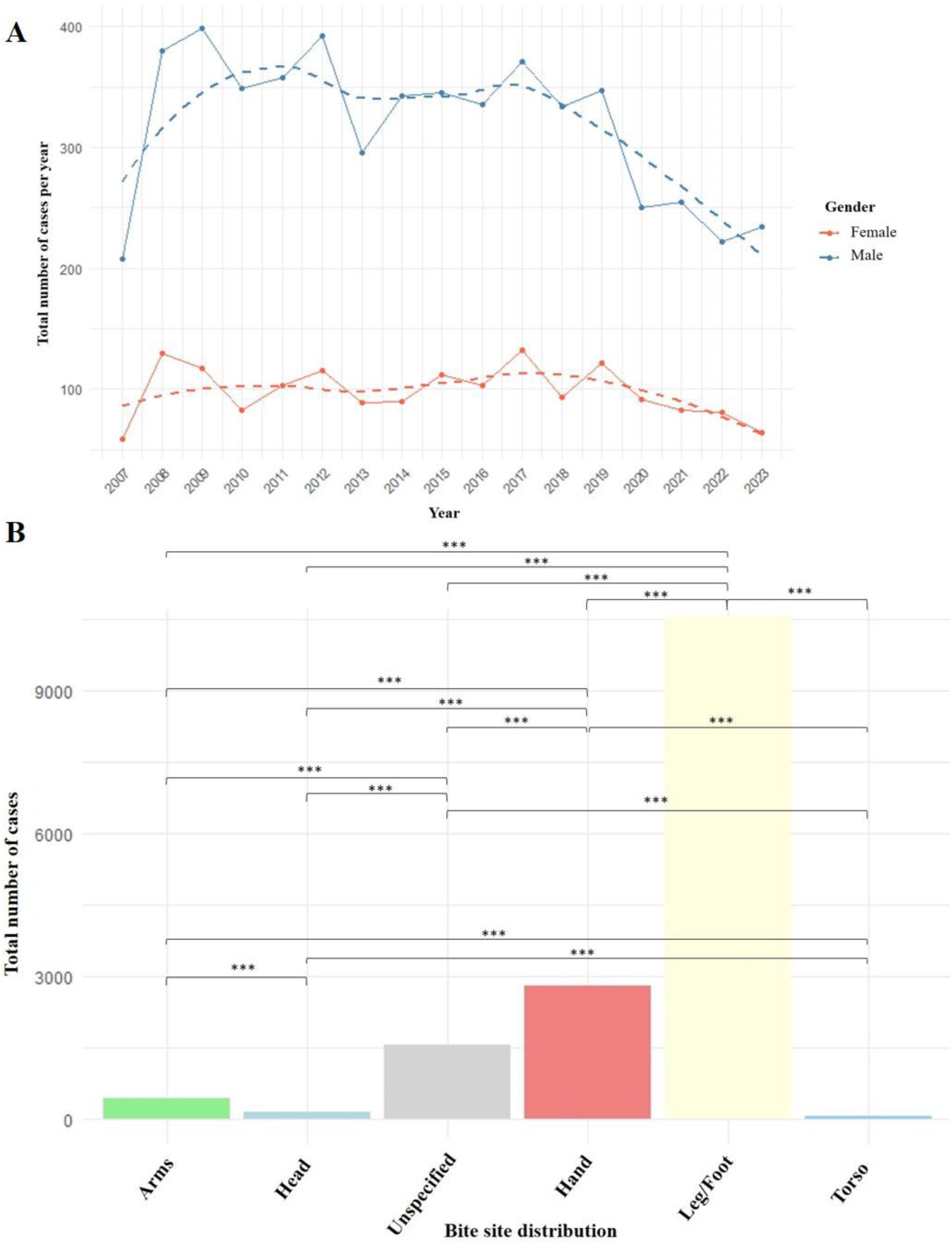
Characterization of snakebite envenomings in Mato Grosso do Sul (2007–2023) by sex and anatomical site of bite. (A) Distribution of cases by sex, showing a strong predominance among males (n = 5,408; 76.5%) compared to females (n = 1,664; 23.5%), an approximate 3:1 ratio (χ² test, p < 0.001). This difference remained consistent over time, reflecting the predominantly male occupational exposure profile, especially in agricultural activities. (B) Percentage distribution by anatomical site, with bites mainly on lower limbs (legs and feet), followed by hands and arms, and rare involvement of trunk and head. The χ² test indicated highly significant differences among proportions (χ² = 31.45; p < 2.2 × 10⁻¹⁶), consistent with exposure patterns typical of rural and fieldwork settings.

Regarding age distribution, most snakebite envenoming occurred among individuals aged 20–39 years (34.7%) and 40–59 years (29.8%), together accounting for over 64% of all cases. Children aged 0–9 years represented 8.8% of cases, whereas older adults (≥60 years) accounted for 11.5%, highlighting their particular vulnerability to complications. These findings indicate that the economically active population constitutes the most exposed group, while also emphasizing the need for special attention to the youngest and oldest individuals, due to their greater susceptibility to severe outcomes. It is worth noting that some variables showed a considerable proportion of missing data, which may limit more detailed analyses. This limitation was more evident during the COVID-19 pandemic, when the average percentage of missing data increased to 6.4%, compared to 4.7% in the pre-pandemic years.

In addition to differences by sex, we also evaluated the anatomical site of the bite. The chi-square test revealed highly significant differences among sites (χ² = 31,450; p < 2.2e–16). Legs and feet were the most affected regions, followed by hands and arms, whereas bites to the trunk and head were rare. Pairwise comparisons confirmed significant differences between all anatomical sites (p < 0.001), except for a few residual proportions (**Table S1**). These results highlight the predominance of lower limbs and hands as the main targets of snakebites (**Figure 7**).

Furthermore, analysis of the time elapsed until medical care showed that most accidents were treated within three hours after the bite, with predominance of the “≤1 hour” and “1–3 hours” categories. The distribution of response times varied significantly both among time intervals and across years (Kruskal–Wallis, χ² = 60.39; df = 16; p < 0.001). These findings indicate that treatment patterns did not remain constant over the study period, reflecting temporal fluctuations in the relative frequency of response times (**Figure 8A**).

**Figure 8.**
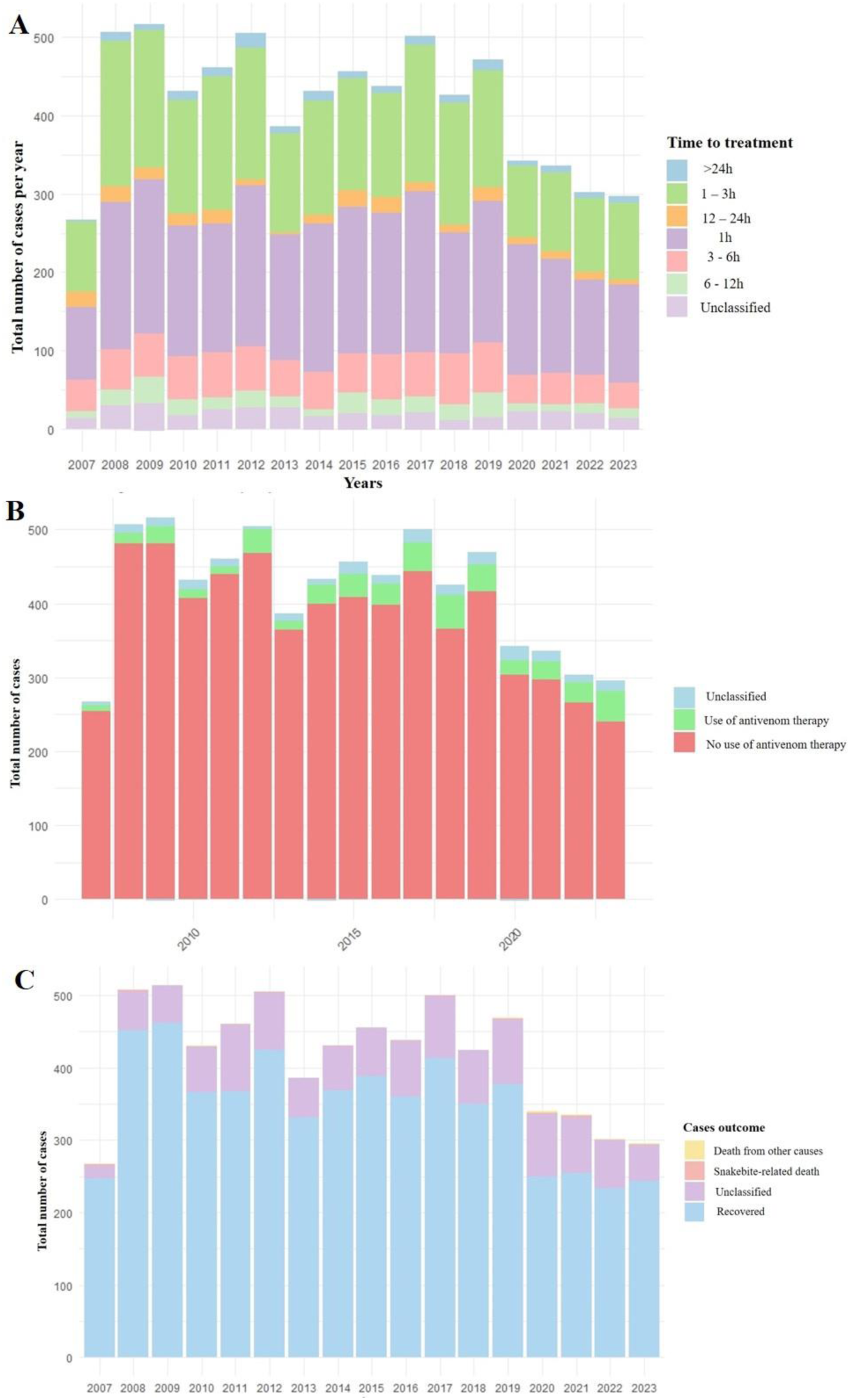
Clinical and healthcare aspects of snakebite envenomings in Mato Grosso do Sul (2007–2023). (A) Distribution of medical care intervals after the bite, showing predominance of treatments within 3 hours. Statistical analysis indicated significant differences across intervals and years (Kruskal–Wallis, χ² = 60.39; df = 16; p < 0.001), suggesting temporal variations in response time. (B) Proportion of cases treated with antivenom, showing broad predominance of treatment (98.2%) compared with cases without recorded therapy (1.8%) (χ² = 2000.8; df = 1; p < 0.001; 95% CI = 95.3–97.7%). Despite minor annual variation (p = 0.0017), therapeutic coverage remained consistently high. (C) Clinical outcomes, with cure in approximately 83% of cases. Among fatalities, two-thirds were directly attributable to the bite and one-third to other causes (p = 0.001). Kruskal–Wallis test indicated significant variation in outcomes across years (p = 0.0006), although cure remained the predominant result.

In addition to analyzing response times, the use of antivenom therapy was also assessed in reported cases. Antivenom administration was predominant, occurring in 98.2% of incidents, whereas only 1.8% of patients did not receive this treatment. A proportions test confirmed a highly significant difference between the two categories (Chi-square = 2000.8; df = 1; p < 0.001; 95% CI for the difference = 95.3–97.7%). Furthermore, the frequency of antivenom use varied significantly across the years (p = 0.0017), although its predominance remained consistent throughout the study period (**Figure 8B**).

Regarding clinical outcomes, favorable evolution was predominant. Most patients recovered, representing approximately 83% of all notifications. Among recorded deaths, around two-thirds were directly attributable to snakebite cases, while one-third were classified as due to other causes, a difference that was statistically significant (p = 0.001). The Kruskal–Wallis test indicated significant temporal differences in clinical outcomes over the years (p = 0.0006), suggesting that although recovery remained the most frequent outcome, relevant interannual variations occurred (**Figure 8C**).

Specifically, recovery was significantly more frequent than all other outcome categories (p < 2.2e–16). In contrast, fatalities—whether directly attributable to snakebite or due to other causes—were rare but differed significantly from each other (p = 0.001), with deaths directly related to snakebite being more common. Annual proportions of deaths directly attributable to snakebite showed considerable fluctuations, with higher values in specific years, such as 2020 (0.59%) and 2021 (0.60%), and no occurrences in others, including 2009, 2013, 2015, and 2022.

Although the total number of deaths was low (n = 18; 0.27%), the age distribution of these cases revealed a notable pattern: 11 deaths occurred in individuals aged 40 years or older, and 4 occurred in children aged 1–4 years. These findings suggest that age extremes constitute particularly vulnerable groups for fatal outcomes in snakebite cases, possibly due to lower physiological reserve, the presence of comorbidities in older adults, and the relatively higher impact of venom in young children. Similar to the distribution of cases by snake genus, 15 of the 18 deaths occurred in men (83.3%), compared to only 3 deaths in women (16.7%). While men accounted for roughly three times more cases overall, the male-to-female ratio among fatalities increased to 5:1, indicating a proportionally higher lethality in men. Of note, five of the 18 deaths (27.7%) occurred in Indigenous individuals, reflecting the high proportion of Indigenous people among snakebite victims. Interestingly, only one of the 18 deceased patients did not receive antivenom therapy. Regarding response time, five deaths occurred in patients attended within 1 hour, 5 between 1 and 3 hours, 1 between 6 and 12 hours, and 4 between 12 and 24 hours, while three cases lacked this information.

Analysis of the association between snakebite cases and occupational activities revealed that only 27.9% of cases were classified as work-related, whereas the majority (72.1%) were not associated with employment. A proportions test indicated that this difference was highly significant (p < 2.2e–16; 95% CI: –0.4528 to –0.4304), confirming that non-occupational accidents predominate in the state. Nonetheless, the fact that nearly one-third of cases were work-related highlights the importance of this component in the epidemiological profile, particularly considering the role of agricultural, extractive, and rural tourism activities in MS.

The analysis of snakebite distribution by biome revealed interesting patterns. Visually, *Bothrops* exhibited a higher relative risk in the Pantanal region, whereas the other genera showed similar risk levels between the Cerrado and Atlantic Forest. Only *Lachesis* displayed a tendency toward lower risk in the Atlantic Forest, albeit minimally (Figure S4A). Statistical testing confirmed the absence of significant differences among biomes for all genera assessed (*Bothrops*: χ² = 5.15, df = 2, p = 0.076; *Crotalus*: χ² = 0.09, p = 0.958; *Micrurus*: χ² = 1.15, p = 0.564; *Lachesis*: χ² = 1.53, p = 0.467). Nevertheless, the observed pattern for *Bothrops* suggests a potentially higher vulnerability in the Pantanal, which may be linked to the environmental characteristics of the region—a hypothesis that warrants further discussion (**Figure S4B**).

Analysis by climate type indicated specific risk patterns. *Crotalus* and *Micrurus* appeared to present higher relative risk in semi-humid and hot climate areas, whereas *Bothrops* and *Lachesis* showed a more homogeneous distribution across the evaluated climatic categories (**Figure S5A**). However, Kruskal–Wallis tests did not detect statistically significant differences among the groups (*Bothrops*: χ² = 0.30, p = 0.586; *Crotalus*: χ² = 1.85, p = 0.173; *Micrurus*: χ² = 1.68, p = 0.195; *Lachesis*: χ² = 0.01, p = 0.932). These findings suggest that although visual patterns associate *Crotalus* and *Micrurus* with semi-humid regions, the magnitude of cases was insufficient to confirm these differences statistically (**Figure S5B**). Exploratory analyses with continuous climatic variables indicated a weak positive association between mean annual temperature and the number of cases recorded (β = 1.35; p = 0.004; R² = 2.6%), while no significant association was observed for precipitation (p = 0.375). In a multiple regression model, temperature remained a significant predictor even after adjustment for precipitation, though the overall explanatory power remained low (R² = 3.5%). These results should be interpreted cautiously given the incomplete nature of the climatic data (**Table S2**).

Considering that most reported snakebite cases were not work-related, we hypothesized that a substantial portion of these events could be associated with leisure activities, particularly tourism. Therefore, we investigated the relationship between municipal tourism classification and snakebite occurrence. Spatial analysis indicated that municipalities classified as tourist destinations had higher absolute numbers of snakebite cases (**Figure S6**), especially those categorized as A and B. These results also revealed a higher relative risk for *Bothrops*, *Crotalus*, and *Micrurus* in tourist municipalities. Statistical analyses confirmed these differences, with Kruskal–Wallis tests showing significant associations between snakebite occurrence and municipal tourism classification for *Bothrops* (χ² = 6.17, p = 0.0457), *Crotalus* (χ² = 7.28, p = 0.0262), *Lachesis* (χ² = 6.71, p = 0.0349), and *Micrurus* (χ² = 6.19, p = 0.0451) (**Figure 9A**).

**Figure 9.**
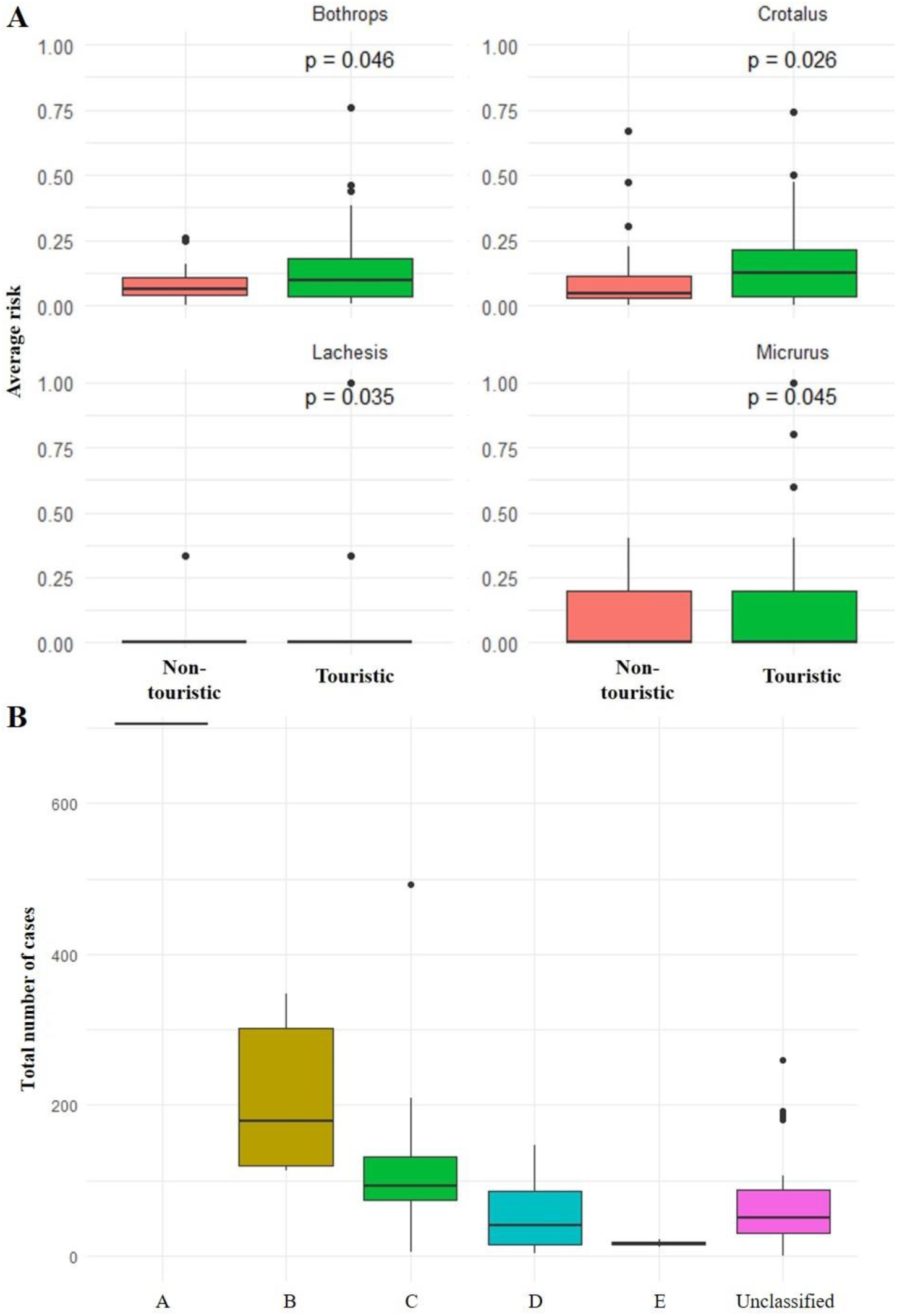
Relationship between municipal tourism classification and snakebite envenomings in Mato Grosso do Sul (2007–2023). (A) Statistical association between snakebite occurrence and tourism classification categories. The Kruskal–Wallis test revealed significant differences for all snake genera — *Bothrops* (χ² = 6.17; p = 0.0457), *Crotalus* (χ² = 7.28; p = 0.0262), *Lachesis* (χ² = 6.71; p = 0.0349), and *Micrurus* (χ² = 6.19; p = 0.0451) — indicating influence of rural and ecotourism activities on exposure risk. (B) Total number of cases by tourism category (A–E and non-classified municipalities). A progressive reduction in cases was observed from highly developed (A–C) to lower-ranked (D–E) categories, with a slight increase among non-classified municipalities. The Kruskal–Wallis test (p < 0.01), followed by Dunn’s test with Bonferroni correction, confirmed significant group differences, and the negative binomial regression model corroborated that less-structured municipalities presented substantially lower risk compared to category A.

When analyzing the relationship between snakebite cases and tourism categories (A–E or uncategorized), a significant association was observed (Kruskal–Wallis χ² = 20.36, p = 0.001), suggesting a decrease in cases as tourism category decreased (**Figure 9B**). However, post-hoc analysis with Bonferroni-adjusted p-values showed that only category B municipalities had significantly higher case numbers compared to category D (p = 0.016) and uncategorized municipalities (p = 0.037). Negative binomial regression confirmed this trend, indicating that municipalities in category A (highest tourism infrastructure) had, on average, the highest number of cases, whereas lower categories exhibited progressively fewer cases. Compared to category A, municipalities classified as D (IRR = 0.08; 95% CI: 0.01–0.30; p = 0.003), E (IRR = 0.02; 95% CI: 0.002–0.18; p < 0.001), and uncategorized (IRR = 0.10; 95% CI: 0.01–0.38; p = 0.007) showed statistically significant reductions in the number of cases.

Considering that part of the snakebite cases is associated with leisure and tourism activities, as explored in the previous section, it is also relevant to investigate accidents related to occupational activities. In this context, we assessed whether different forms of land use and occupation could influence the occurrence of snakebites, given that agricultural areas, pastures, or even urban environments may reflect distinct patterns of human exposure to snakes (**Figure S7A**).

The analysis of the relationship between land use and snakebite incidence revealed significant associations for some occupation types. Spearman correlation analysis indicated that several land use variables were moderately to strongly correlated with the total number of snakebite cases. The strongest positive correlations were observed for total municipal area (ρ = 0.60), artificial areas (ρ = 0.55), mosaics of grassland areas (ρ = 0.51), and forested vegetation (ρ = 0.48). Agricultural areas (ρ = 0.31), pastures (ρ = 0.38), and grassland areas (ρ = 0.33) showed weaker but still significant positive correlations (p < 0.001). In contrast, the presence of inland water bodies showed no relevant relationship with snakebite occurrence (ρ ≈ 0) (**Figure S7B**). Multiple regression modeling considering different land use categories explained approximately 83% of the variation in snakebite cases among municipalities (R² = 0.83; p < 0.001). Coefficients indicated positive associations for urban areas (β = 2.39; p < 0.001), agricultural areas (β = 0.027; p = 0.0006), and grassland areas (β = 0.044; p < 0.001). In contrast, wetlands (β = -0.281; p < 0.001) and silviculture areas (β = -0.037; p = 0.028) showed negative associations. Other variables, such as forests and mosaics, did not exhibit significant effects.

When integrating the tourism and land use variables into a combined model, the association with snakebite cases remained consistent. The model including both tourism and land use showed comparable or slightly improved performance (AIC = 833.5) relative to the global model that also incorporated biome and climate variables (AIC = 835.2), suggesting that the latter did not provide additional explanatory power. Among the analyzed factors, agricultural (p = 0.007) and pasture areas (p = 0.029) remained positively associated with the number of cases, whereas artificial areas exhibited only a marginal association (p = 0.084). In turn, tourism categories presented high but non-significant coefficients when considered jointly with land use, indicating overlapping effects. Overall, these results reinforce that land occupation, particularly agriculture and pastures, and the presence of tourist flows are the main factors associated with the spatial variation of snakebite cases, whereas biome and climate contributed little to explaining the observed patterns (**Table S3**).

## Discussion

Although the state of Mato Grosso do Sul recorded around 7,000 snakebite cases between 2007 and 2023, this figure represents only 1.7% of the 418,383 cases reported in Brazil during the same period. These numbers hold local epidemiological relevance, as they reflect the overlap between human populations and ecosystems with high snake diversity, particularly within the Cerrado, Atlantic Forest, and Pantanal biomes, which are widely distributed across the state [**18**].

The relatively small contribution of MS to the national total does not diminish the importance of the problem at the state level, since regions with fewer absolute cases may still present focal areas of risk and particularly vulnerable populations. These conditions pose challenges for surveillance and access to specific medical care. Such local findings are consistent with the national pattern of a high burden of snakebite cases in tropical rural areas, and with previous reviews identifying *Bothrops* as the main genus responsible for envenomations in Latin America and Brazil [**18, 29, 30**].

According to the World Health Organization guidelines, it is essential to prioritize efforts and resources in areas where snakebites persist, even when the state does not rank among those with the highest absolute numbers of notifications [**1**]. A detailed understanding of local epidemiology is crucial, since changes in surveillance practices, access to healthcare services, and educational campaigns can affect temporal trends and inter-state comparisons. Furthermore, risk-modulating factors, such as land use and transformation, human mobility, and the distribution of venomous snake species, may influence snakebite occurrence directly or indirectly, reinforcing the need for continuous monitoring to reduce morbidity and mortality due to snake envenoming [**1**]

Between 2007 and 2023, annual variation in snakebite incidents was observed in MS, without a significant linear trend, as indicated by the fitted regression and ARIMA models. Non-parametric Kruskal–Wallis and Dunn tests also did not reveal statistically significant differences between years, suggesting relative stability in the time series. This consistency may reflect a consolidated endemic pattern, in which structural, climatic, and sociodemographic factors maintain a constant risk over the years, with minor seasonal fluctuations. These findings are corroborated by other regional studies in Brazil, which reported stability or a slightly increasing trend over the last two decades, mainly attributed to the maintenance of agricultural activities and population growth in rural areas [**9,13,14,17,21,30,31,32,33**]. At the national level, annual fluctuations without robust statistical trends have also been observed, reinforcing that interannual variation is more likely driven by environmental and operational conditions (rainfall, surveillance efforts) than by real changes in incidence [**30**].

The absence of a significant linear trend in the ARIMA model indicates that, despite annual fluctuations, the average number of cases remained relatively constant over time. This pattern is consistent with the hypothesis of stable endemicity, characteristic of regions where envenoming risk is persistently maintained by favorable ecological and socioeconomic contexts [**2,29,34**]. The slight increases observed in some years may be attributed to local climatic variations, particularly increased precipitation, climatic anomalies (El Niño/La Niña), or temporary intensification of epidemiological surveillance [**21**].

It is noteworthy that years coinciding with La Niña events corresponded to peaks in snakebite cases in MS. A sharp increase was observed in 2008–2009, corresponding to the 2007–2009 La Niña event; another rise occurred in 2011–2012, associated with the 2010–2012 event; and a further increase in 2017, coinciding with the 2016–2018 episode. Only during the 2020–2023 La Niña event was this association not observed, a period marked by a slight reduction in cases. The decrease in cases from 2020–2022 may be related to reduced exposure during the COVID-19 pandemic and/or underreporting, consistent with national findings showing a decline in notifications of venomous animal incidents in Brazil after 2020 (APC: –0.77; p<0.043) [**35, 36**]. Although this association is purely observational, climatic variations linked to ENSO phenomena (El Niño/La Niña) have previously been associated with changes in snakebite incidence in other tropical regions, suggesting a possible environmental influence on local risk [**37**].

The monthly analysis revealed a recurring seasonal pattern, with the highest number of cases occurring between January and April, coinciding with the rainy season in Mato Grosso do Sul. The SARIMA model confirmed this periodicity, indicating a stable temporal structure of annual peaks at the beginning of the year. Although the overlap with the rainfall regime is evident, regression analyses did not identify a statistically significant association between precipitation and the number of cases (β = 0.0017; p = 0.375; R² = 0.25%), possibly due to incomplete rainfall data and spatial heterogeneity of meteorological stations. On the other hand, average temperature showed a positive, albeit weak, association with accidents (β = 1.35; p = 0.004; R² = 2.6%), suggesting that temperature variations may have a more consistent influence on both snake and human activity. Thus, the observed seasonality appears to reflect the combined effects of environmental and behavioral factors, rather than a direct linear response to rainfall. This pattern has also been reported in other tropical regions, where warmer and more humid periods favor increased snake activity and heightened risk of human contact [**29, 30**].

The distribution of venomous snakebites in MS maintained the classic epidemiological pattern observed in Brazil, with a predominance of cases among males and young adults. These groups engage in the primary risk activities, ranging from soil management, livestock handling, and vegetation clearing, to ecotourism and fishing, which are highly prevalent in MS and appear to facilitate snake-human contact in rural and peri-urban environments [**14,18,30,32**]. The predominant age group, between 20 and 49 years, corresponds to the segment with the highest productive activity and mobility in rural areas, aligning with findings from national and Latin American studies. Although less frequent, accidents in children and older adults are epidemiologically relevant due to their association with greater severity and lethality, resulting from lower body mass and the presence of comorbidities [**29,38,39**].

The spatial distribution of snakebite incidents in MS showed marked heterogeneity among municipalities, with the highest concentration of cases in the Central-North and Pantanal regions, particularly in municipalities such as Corumbá, Aquidauana, Coxim, Miranda, and Campo Grande, as evidenced in Figures 3, 4, and 5. These municipalities encompass transition areas between the Cerrado and Pantanal biomes, characterized by high snake diversity and intense interfaces between natural environments and agricultural activities [18]. The northern part of the state, especially areas associated with the Taquari and Miranda river basins, exhibited higher incidence rates, possibly related to dispersed rural occupation, high density of *Bothrops* snakes, and limited rapid access to healthcare services. This pattern is similar to that described in other states of the Central-West and Northern Brazil, where human contact with native vegetation and extensive livestock areas is the main risk factor [**14**].

However, when considering relative risk by snake genus (Figure 6) within each municipality, a differentiated distribution is observed, reflecting local ecological variations and specific human exposure patterns. *Bothrops* accidents were the most frequent in the state, primarily concentrated in Campo Grande, Aquidauana, Corumbá, and Dourados, consistent with the wide distribution of this genus in Cerrado environments and anthropized areas [**14,18,40**]. For the genus *Crotalus*, although the absolute number of cases is lower, municipal relative risk was high in Campo Grande, Dourados, and Antônio João, indicating a higher proportion of rattlesnake incidents in these locations, possibly associated with the presence of open areas and pastures, the preferred habitats of *Crotalus* [**40**]. *Micrurus* accidents showed a higher proportion in Dourados, Sidrolândia, and Caarapó, followed by Campo Grande and Corumbá, although absolute incidence remained low. These findings suggest that coral snake risk, while low in frequency, is spatially focused and possibly related to the presence of forest fragments and sandy soils, which are favorable habitats for fossorial species [**41,42**]. Finally, the municipality of Bonito stands out for *Lachesis*, where, despite the low absolute number of cases, most reported accidents were attributed to this genus, increasing its proportional risk. This pattern highlights the importance of considering not only total incidence but also the genus-specific composition of accidents in each locality, allowing the identification of differentiated risk zones and the optimization of surveillance strategies and allocation of specific antivenoms [**1**].

In addition to differences by sex and age group, the distribution of snakebite cases by anatomical site was evaluated, revealing a statistically robust pattern with highly significant differences in the proportion of bites by body segment (χ² = 31,450; p < 2.2 × 10⁻¹⁶). Legs and feet were the most affected sites, followed by hands and arms, while the trunk and head presented much lower frequencies (**Figure 7**). This anatomical pattern reflects the most common mode of exposure, in which accidental contact with snakes occurs during activities in rural areas, movement through vegetation, or handling soil, situations in which the lower limbs are most vulnerable [**14, 43**]. The high frequency of bites to the hands and arms underscores the risk associated with attempts to capture or handle the animal, a practice reported in national and Latin American studies [**29, 43**].

Regarding the time elapsed until medical care, most cases were attended within three hours of the incident, a favorable pattern, as national literature indicates that treatment administered six hours or later is associated with greater severity of envenomation (odds ratio for moderate to severe cases ranging from 1.25 to 3.03 depending on the snake genus) [**44**]. Reviews on the topic corroborate that delays in antivenom administration increase the risk of severe complications, amputations, tissue necrosis, and death [**45**]. Recent Brazilian studies highlight that three hours may represent a critical window to reduce the likelihood of adverse outcomes, emphasizing the importance of a coordinated snakebite care network with optimized transport and treatment flows [**8**]. Antivenom therapy was administered in 98.2% of cases, including the majority of patients with severe progression, supporting findings that rapid access to treatment and antivenom use can substantially reduce severity and mortality [**32**].

Cases showed a favorable progression, with approximately 83% resulting in recovery. Among the deaths (18 – 0.27%) recorded during the study period, five (27.7%) occurred in self-declared Indigenous individuals, a proportion that is striking when considering that 16.3% of all reported cases also involved Indigenous people—a disproportionately higher representation compared to their presence in the state. This overrepresentation suggests greater epidemiological and social vulnerability, likely related to proximity to rural and forested areas, subsistence activities, and barriers to timely and specialized medical care [**11,32**]. MS stands out nationally for hosting one of the largest Indigenous populations in Brazil, and, proportionally, the frequency of snakebite envenoming among Indigenous individuals is only surpassed by the states of Amazonas and Roraima, which similarly exhibit extensive overlap between traditional territories and areas of snake occurrence [**11,33**].

Additionally, among the 18 recorded deaths, 11 occurred in individuals over 40 years of age, and 4 in children aged 1–4 years, highlighting greater vulnerability at both ends of the age spectrum. This age distribution is consistent with the literature, which recognizes children and the elderly as groups at higher risk of adverse outcomes [**29,32,34**]. In children, lower body mass and a higher venom-to-weight ratio result in greater clinical severity and increased risk of shock, renal failure, and extensive hemorrhage, even in cases involving relatively low venom doses [**33,38**]. Moreover, immature immunological and metabolic responses, along with difficulties in communication and early diagnosis, contribute to worse prognosis [**46**]. In older adults, factors such as pre-existing comorbidities (hypertension, diabetes, chronic kidney disease), reduced inflammatory response, and the use of interfering medications (anticoagulant, antihypertensive) may exacerbate systemic effects of envenomation, increasing mortality and recovery time [**32**]. In both groups, delays in medical care and limited access to facilities with antivenom further worsen prognosis.

When evaluating the cases that progressed to death, there was considerable variation in time to medical care: five deaths occurred in patients attended within 1 hour, five between 1–3 hours, and four between 12–24 hours. It is also noteworthy that only one of the recorded deaths did not receive antivenom therapy. These data indicate that, although prompt medical attention and the use of antivenom are crucial factors for reducing morbidity and mortality, they are not the sole determinants of clinical outcomes. Even among patients who received early care, death may occur due to a high venom load, bites in highly vascularized body regions, or individual conditions that increase vulnerability, such as extreme age (children and elderly), pre-existing comorbidities (cardiovascular disease, diabetes, renal insufficiency), or an exaggerated inflammatory response [**32,38,47**].

Projections of snakebite envenoming for the period 2024–2026 indicate a tendency for case numbers to remain stable in municipalities historically most affected, such as Campo Grande, Aquidauana, Corumbá, and Dourados (**Figure S1**). In contrast, Sidrolândia is projected to experience a slight increase in cases, possibly associated with agricultural expansion. These locations combine higher human population density with extensive rural–urban interface areas and biomes of high snake diversity, factors that sustain a constant risk of exposure [**18,30**]. ARIMA models applied to municipal time series indicate overall stability with moderate seasonal fluctuations and no marked upward trend, a pattern consistent with national stabilization observed from 2017 onwards. Complementarily, the SARIMA model, incorporating monthly seasonality, confirmed that peak months remain between January and April, reflecting the strong influence of the rainy season and increased agricultural activity on snakebite occurrence [**18,32,37,48**]. This pattern reinforces that, despite interannual variation, the seasonal trend tends to remain stable, with higher risk concentrated at the beginning of the year.

It should be noted, however, that forecast uncertainty increases as projections extend further from the historical series, particularly in municipalities with low numbers of notifications. Therefore, results should be interpreted as indicative, serving as a support tool for planning surveillance actions and antivenom distribution according to predicted seasonality and spatial risk profiles. Finally, future analytical models could integrate climatic and environmental variables, such as precipitation, temperature, land use and cover, and vegetation indices, which could potentially improve predictive accuracy and better elucidate the influence of ecological factors on the dynamics of snakebite envenoming at the municipal level.

The analysis of snakebite distribution by biome revealed distinct risk patterns among snake genera, although without statistically significant differences. Visually, *Bothrops* exhibited higher relative risk in the Pantanal region, whereas *Crotalus* and *Micrurus* showed similar distributions between the Cerrado and Atlantic Forest. *Lachesis* showed a tendency toward lower risk in the Atlantic Forest, although this pattern was not pronounced (**Figure S4A**).

Confirmatory statistical analysis (chi-square test) did not identify significant differences between biomes, suggesting relative homogeneity in snakebite distribution across the state’s main environments. However, the observed pattern for *Bothrops*, with predominance in the Pantanal, is epidemiologically relevant, as it may reflect specific ecological and occupational characteristics of this biome, including high wildlife density, extensive livestock activities, and seasonal flooding, which facilitate movement of both snakes and human populations [**18,49**]. Additionally, the environmental and climatic heterogeneity of the Pantanal, combined with its connectivity to Cerrado and Atlantic Forest areas, creates complex ecological mosaics that hinder the delineation of fixed risk zones and partly explain the lack of statistical significance in comparative tests [**50**].

Analysis according to climatic types indicated specific risk patterns. Visually, *Crotalus* and *Micrurus* showed higher relative risk in semi-humid and warm climates, whereas *Bothrops* and *Lachesis* exhibited a more homogeneous distribution across the evaluated climatic categories. However, Kruskal–Wallis tests did not detect significant differences between groups, suggesting that, although visual association patterns exist, the magnitude of the records was insufficient to statistically confirm these differences. Complementary analyses using continuous variables showed a positive, albeit weak, association between mean annual temperature and the number of reported cases, whereas precipitation did not have a significant effect. Even in multiple adjusted models, temperature remained statistically significant but with low explanatory power, indicating that climate acts as a modulating factor rather than a determinant of snakebite envenoming. These findings are consistent with previous studies linking higher temperatures to increased activity of snakes and their hosts, as well as greater human exposure during rural and recreational activities [**32,37**].

On the other hand, periods of heavy rainfall and flooding, characteristic of the Pantanal climate and parts of the southern Cerrado in MS, may both increase snake displacement into inhabited areas and reduce case reporting due to lower human mobility [**31,51**]. The lack of a strong association with precipitation observed in this study may reflect regional microclimatic heterogeneity and the combined effect of multiple factors, including land use, agricultural seasonality, and altitude, which influence both snake ecology and human exposure. Thus, although climatic conditions are recognized to affect snake ecology and behavior, their direct relationship with snakebite envenoming should be interpreted within a broader, multicausal, and spatially variable context.

Although the majority of recorded snakebite envenoming were not directly associated with occupational activities, spatial analysis showed that municipalities with a high tourism classification (categories A and B of the Brazilian Tourism Map) concentrated the highest number of cases. These findings suggest that the flow of visitors and the intensification of recreational activities in natural environments, such as trails, rivers (fishing), and dense vegetation areas, may increase exposure to snakes for both local residents and tourists. The significant association identified between the occurrence of snakebites and tourism classification supports this hypothesis.

Although statistically significant differences were observed only between municipalities, in category B and those classified as D or uncategorized, the visual pattern indicates a general trend of decreasing cases with lower tourism categories. It is also noteworthy that uncategorized municipalities appear, in terms of case numbers, at levels between categories D and E. Importantly; the absence of an official classification does not necessarily imply the absence of tourism activity but may reflect a lack of formal registration or updating in the Brazilian Tourism Map. Thus, some of these municipalities may have relevant tourist flow that is not yet institutionally recognized. The decreasing incidence pattern with lower tourism classification suggests that the presence of well-established tourism infrastructure can paradoxically represent an increased risk factor, particularly when activities involve direct contact with natural environments, such as ecotourism, fishing, and adventure sports [**52**].

In a study conducted in France, a strong association was observed between tourism intensity and the increased risk of snakebite envenoming. Municipalities with higher tourism density had significantly greater odds of recording at least one bite over a ten-year period. Localities with 11–20 beds per square kilometer had an odds ratio (OR) of 3.79, while those with more than 100 beds/km² had an OR of 4.56 compared to municipalities with up to 10 beds/km². Similarly, an increase in tourism activity, expressed as the number of beds per 100 inhabitants, was also positively associated with bite risk: municipalities with more than 200 beds per 100 inhabitants had an OR of 4.37 relative to those with fewer than 50. In addition to tourism-related factors, the study found that warmer and sunnier regions had a higher likelihood of snakebites, reinforcing the combined role of climate and human activity in the spatial distribution of snakebite envenoming [**53**].

It is important, however, to interpret these results with caution. The higher number of records in tourist municipalities may reflect not only greater exposure but also better surveillance and reporting capacity, as these localities tend to have improved access to healthcare services and hospital infrastructure, which increases the likelihood of formal event registration [36]. Thus, part of the observed difference may result from observation effects rather than a truly higher incidence. On the other hand, seasonal tourism, with increased population movement during warm and rainy months, may overlap with periods of heightened snake activity, reinforcing the risk during certain times of the year.

The results also indicated that variables associated with land use and occupation were strongly related to the number of snakebite envenoming recorded in the municipalities of MS. The strongest positive correlations were observed for urbanized, agricultural, and pasture areas, suggesting that the risk is not limited to natural environments but also emerges in contexts of rural-urban transition, where there is greater interface between human activities and snake habitats.

This relationship is consistent with patterns observed in other tropical regions, where agricultural expansion, deforestation, and forest fragmentation modify habitats and alter the ecological dynamics of species, bringing them closer to populated areas [**11,12,37,54**]. In particular, the genus *Bothrops*, responsible for the majority of incidents in the state, is highly adaptable to open areas, plantations, and forest edges, which explains its higher occurrence in agricultural and pasture regions [**10,54**]. On the other hand, the positive association with artificial (urban) areas should be interpreted cautiously. Although unplanned urbanization can generate favorable microenvironments (vacant lots, vegetation debris, and rodent presence), part of this pattern may reflect better surveillance and reporting infrastructure. More populous municipalities, with extensive hospital networks and reference facilities, tend to record and report cases more efficiently, which can inflate the absolute number of occurrences without necessarily representing a higher real risk [**36**]. Conversely, the negative associations observed for wetlands and silviculture suggest that these environments function as areas of lower human exposure or have conditions less favorable for encounters between snakes and people. This inverse relationship has been described in studies from the Amazon, where flood-prone areas act as temporary ecological barriers, reducing human traffic and, consequently, the risk of snakebites [**31**].

The multiple regression model indicated that land-use variables explained up to 83% of the spatial variation in snakebite envenoming, outperforming models that included biome and climate, which reinforces that land occupation and use patterns are key determinants of accident distribution in the state. When combined with tourism variables, a plausible overlap of effects was observed, as tourist municipalities often coincide with mixed-use areas, combining agriculture, natural areas, and urban expansion. Thus, integrating different models and analyses highlights that the occurrence of snakebite envenoming in MS results from a complex interaction of environmental, occupational, and socio-spatial factors. Although biomes and climate exert contextual influence, the most robust determinants were land use and occupation, along with variables related to human mobility and tourism. This combination of factors underscores the multifactorial nature of snakebite risk, in which exposure depends not only on the presence of snakes but also on how territories are occupied, exploited, and transformed [**18,54**].

The findings suggest that urban and agricultural expansion, combined with increased tourist flows, tends to amplify human-snake contact, particularly in medium- and large-sized municipalities, which concentrate both diverse economic activities and better surveillance and healthcare infrastructure. This scenario underscores the need for public health strategies integrating epidemiological surveillance, territorial planning, and environmental education, as recommended by the World Health Organization [**1**]. Therefore, surveillance should prioritize municipalities with persistent cases, including those not among the highest in absolute numbers but exhibiting high proportional rates or elevated relative risk, especially among vulnerable populations such as children and indigenous peoples. These groups require tailored care protocols and targeted educational campaigns, taking into account geographic and cultural barriers that hinder access to treatment [**10,35**].

The incorporation of predictive models, such as time-series approaches (SARIMA) and spatial regressions, demonstrates potential to enhance continuous monitoring and anticipate periods and areas of higher risk, supporting seasonal prevention, antivenom distribution, and local team training. Integrating these models into state surveillance routines could strengthen responses to climatic and environmental variations. However, this study has limitations. Data incompleteness and heterogeneity, particularly regarding climate and land-cover variables, may have reduced the explanatory power of some models and limited spatial analysis precision. Underreporting and variability in record quality across municipalities and years are additional sources of potential bias. Furthermore, as an ecological study, observed associations do not imply direct causality and should be explored in future local or individual-level investigations. Despite these constraints, the findings underscore the need for territorially oriented, intersectoral, and context-sensitive snakebite surveillance to reduce morbidity and mortality and support effective, equitable public health responses [**55,56**].

## Conclusion

This study revealed that, between 2007 and 2023, snakebite envenoming in MS remained relatively stable, with an annual average of approximately 415 cases, despite interannual fluctuations and a decrease observed during the COVID-19 pandemic. Temporal analysis showed no consistent directional trend, indicating that incidents follow a stochastic pattern with a strong seasonal component, occurring primarily between January and April. The genus *Bothrops* was the main contributor to snakebite envenoming, showing a broad geographic distribution and predominance in medium- and large-sized municipalities, whereas *Crotalus*, *Micrurus*, and *Lachesis* exhibited lower frequency and more restricted spatial distribution. Spatial analysis revealed significant intra-state heterogeneity, with persistent high-incidence areas in the central-southern and northwestern regions of the state. Cluster analysis and risk mapping identified high-risk municipalities, such as Campo Grande, Aquidauana, and Bonito, highlighting the need for targeted local prevention strategies.

Sociodemographic data indicated a predominance of snakebites among men, mixed-race individuals, and economically active adults, although young children and the elderly exhibited greater vulnerability to severe outcomes. Lower limbs were the most affected anatomical sites, and rapid medical care was common, reflecting the effectiveness of healthcare services in administering antivenom, which was applied in over 98% of cases. Mortality was low, concentrated among extreme age groups and men, including Indigenous individuals, emphasizing the need for attention to vulnerable populations. Analysis of environmental and land-use factors showed that agricultural areas, pastures, and urbanized zones were associated with higher snakebite risk, whereas biome and climate had a limited impact. Municipalities with higher tourist influx also exhibited increased incidence, suggesting overlap between recreational activities, human exposure, and case reporting.

In summary, these results indicate that the distribution and occurrence of snakebite envenoming in MS are primarily determined by local socio-environmental factors, including land use, population profile, and human activities, with a strong seasonal component and spatial heterogeneity. These findings provide essential support for the design of prevention strategies, health resource planning, and the development of public policies aimed at mitigating the risk of snakebite envenoming, particularly in municipalities with greater vulnerability. These findings emphasize the need for integrated environmental and epidemiological monitoring in snakebite surveillance programs.

## Data Availability

All data produced in the present work are contained within the manuscript. Additional data supporting the findings of this study are available from the corresponding author upon reasonable request.

## Declarations

### Ethics approval and consent to participate

Not applicable. This study used anonymized, secondary, publicly available data. Therefore, submission to a Research Ethics Committee was not required, in accordance with Resolution CNS No. 510/2016, which waives ethical review for studies using public-domain information.

### Consent for publication

Not applicable.

### Availability of data and materials

The datasets analyzed in this study are publicly available from the following sources:

- Brazilian Ministry of Health (SINAN) – https://www.dadosus.saude.gov.br/acesso-a-informacao/doencas-e-agravos-de-notificacao-de-2007-em-diante-sinan/
- Brazilian Institute of Geography and Statistics (IBGE) – https://www.ibge.gov.br
- National Indian Foundation (FUNAI) – https://www.gov.br/funai/pt-br/atuacao/terras-indigenas/geoprocessamento-e-mapas
- MapBiomas Project – https://brasil.mapbiomas.org/map/colecao-8/
- National Institute of Meteorology (INMET) – https://portal.inmet.gov.br
- Mato Grosso do Sul Tourism Foundation (FundTur-MS) – https://www.observatorioturismo.ms.gov.br/wp-content/uploads/2022/03/ANUARIO_2021_ BASE2020_VF.pdf

All processed data and analytical codes are available from the corresponding author upon reasonable request.

### Authors’ contributions

Machado MS, Machado AM, and Machado ARSR collected and processed the data.

Machado AM, Machado ARSR, Silva AV, Oda JY, performed the statistical and spatial analyses.

All authors actively participated in the interpretation of results and in writing the manuscript.

All authors read and approved the final version of the paper.

## Acknowledgements

The authors acknowledge the availability of open public data provided by Brazilian institutions, which made this research possible.

---

**Figure S1.**
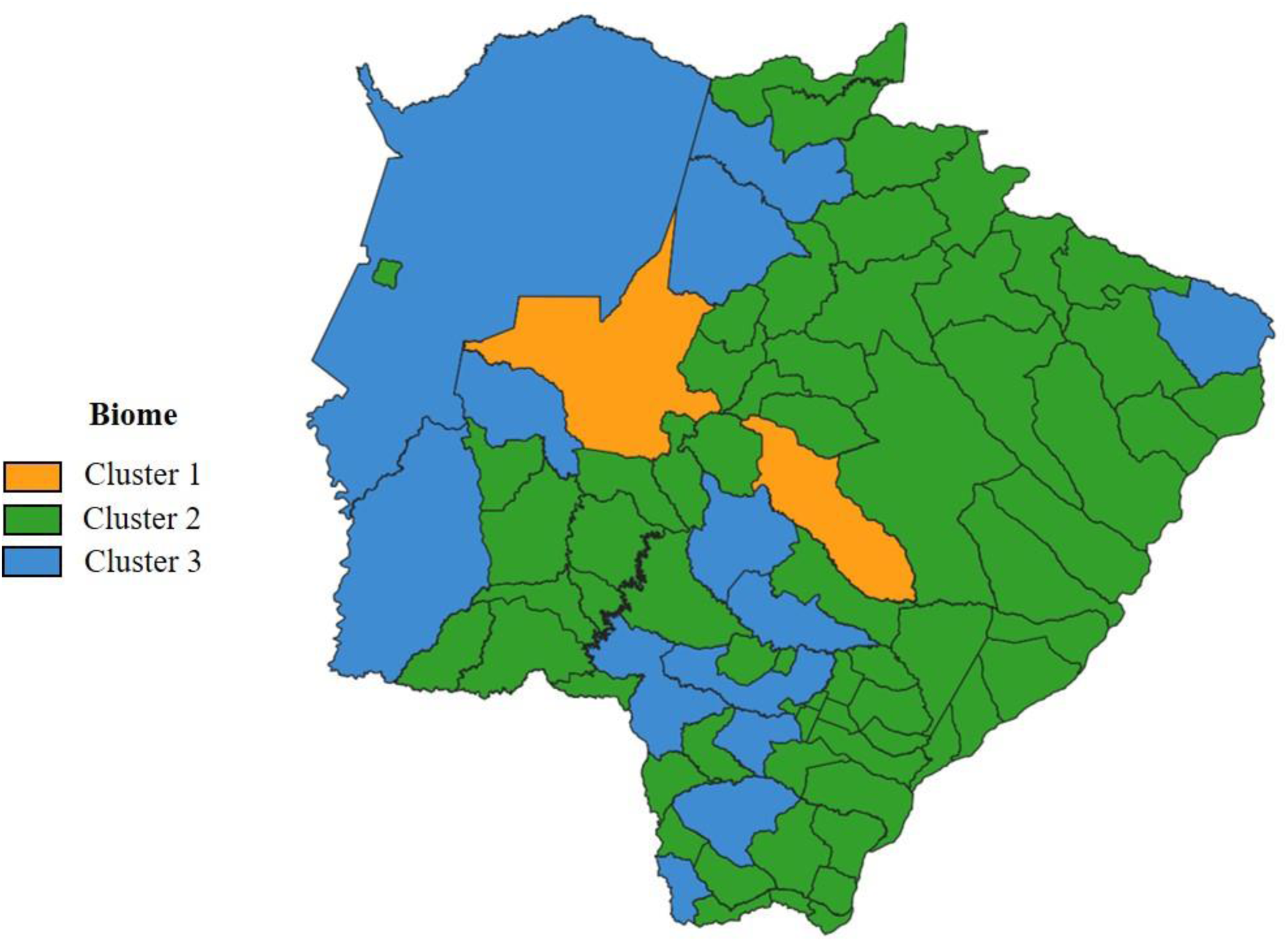
Hierarchical cluster analysis of municipalities in Mato Grosso do Sul based on the total number of snakebite envenomings (2007–2023). Three distinct municipal clusters were identified. **Cluster 1** included high-incidence municipalities such as Campo Grande (705 cases) and Aquidauana (492). **Cluster 2**, the largest, comprised 66 municipalities with low-to-moderate occurrence, ranging from few records (e.g., Anastácio – 4 cases; Anaurilândia – 19) to over 100 (e.g., Bonito – 120; Nova Andradina – 93). **Cluster 3** grouped 11 regional centers, including Dourados (347), Corumbá (302), Sidrolândia (260), and Coxim (193), showing intermediate-to-high incidence. These clusters highlight intra-state heterogeneity in case distribution.

**Figure S1.**
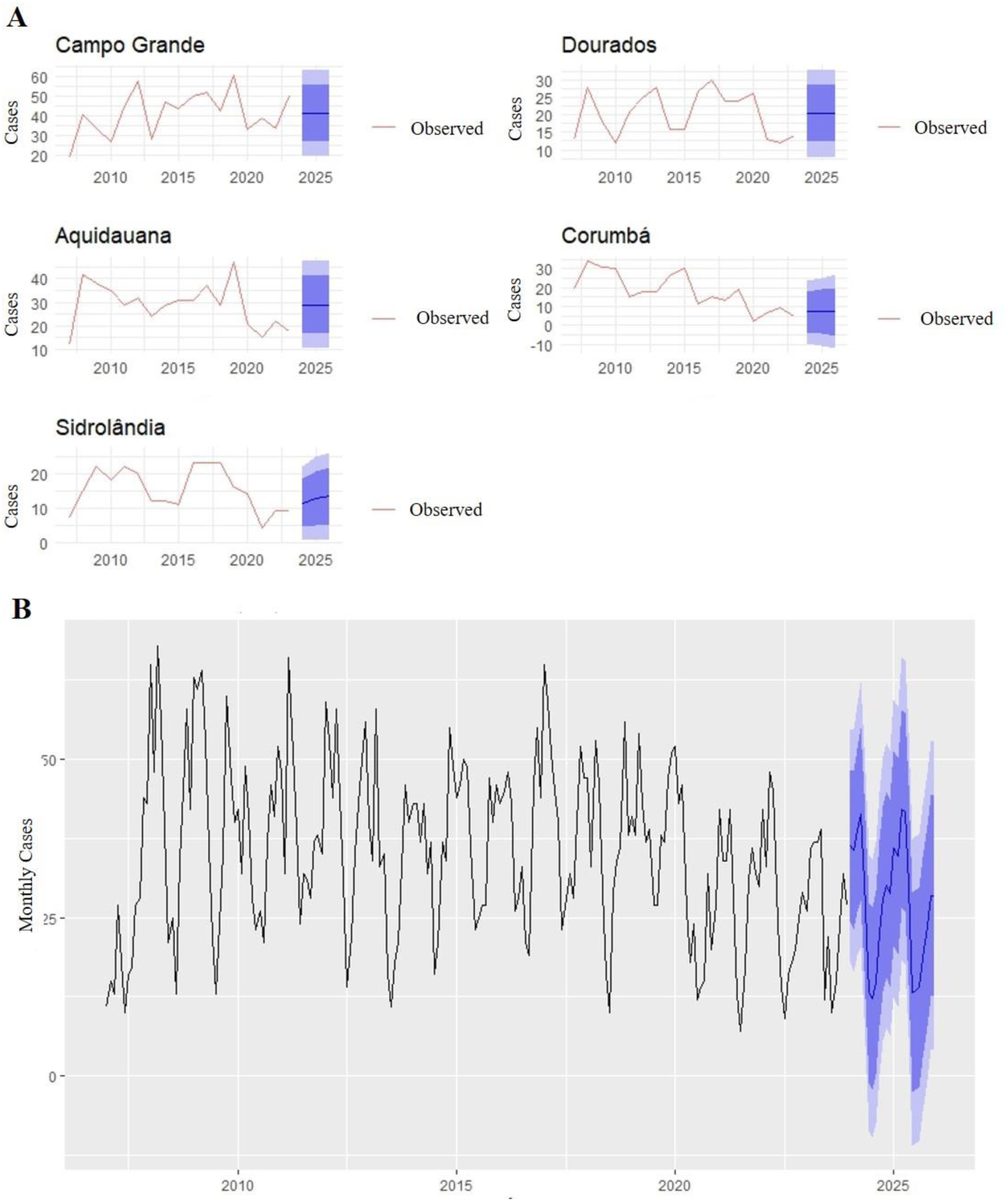
Time-series modeling and forecasts of snakebite envenomings in Mato Grosso do Sul. **(A)** (A) ARIMA models fitted to the five municipalities with the highest number of cases (Campo Grande, Aquidauana, Dourados, Corumbá, and Sidrolândia). Solid lines represent observed data (2007–2023), dashed lines represent forecasts for 2024–2026, and shaded areas indicate 95% confidence intervals. Overall stability is projected, with a slight increasing trend only in Sidrolândia. (B) State-level seasonal forecast for 2024–2025 using a SARIMA model. The curve shows observed monthly values (2007–2023, black line) and projections for 2024–2025 (blue line) with 80% and 95% confidence intervals (shaded areas). The SARIMA (3,0,0)(2,1,0)[12] model indicated maintenance of the typical seasonal pattern, with higher incidence expected during the rainy months (January–April) and reduction during the dry season (June–August).

**Figure S3.**
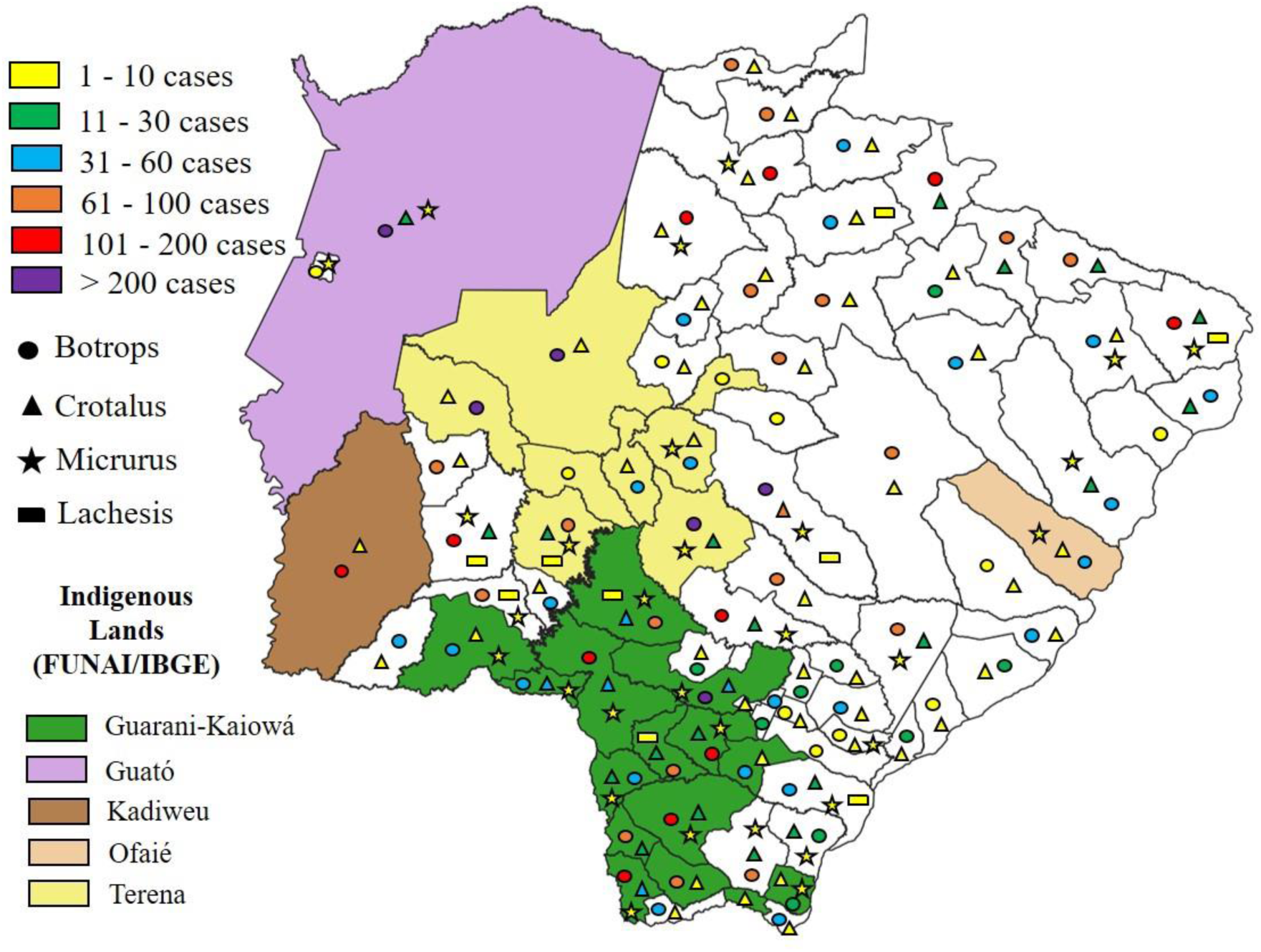
Spatial distribution of snakebite envenomings in Mato Grosso do Sul (2007–2023) overlaid with officially recognized Indigenous Lands. Partial overlap is observed between areas of high incidence and territories occupied by Indigenous peoples such as Guarani-Kaiowá, Guató, Kadiwéu, Ofaié, and Terena. This spatial pattern may help explain the high proportion of cases among self-declared Indigenous individuals (16.3%), placing the state among the three highest nationwide, after Roraima and Amazonas. Results underscore the importance of considering territorial and healthcare-access factors when designing public-health policies for snakebite surveillance and management. (Map based on public data from FUNAI and SINAN/DATASUS.)

**Figure S4.**
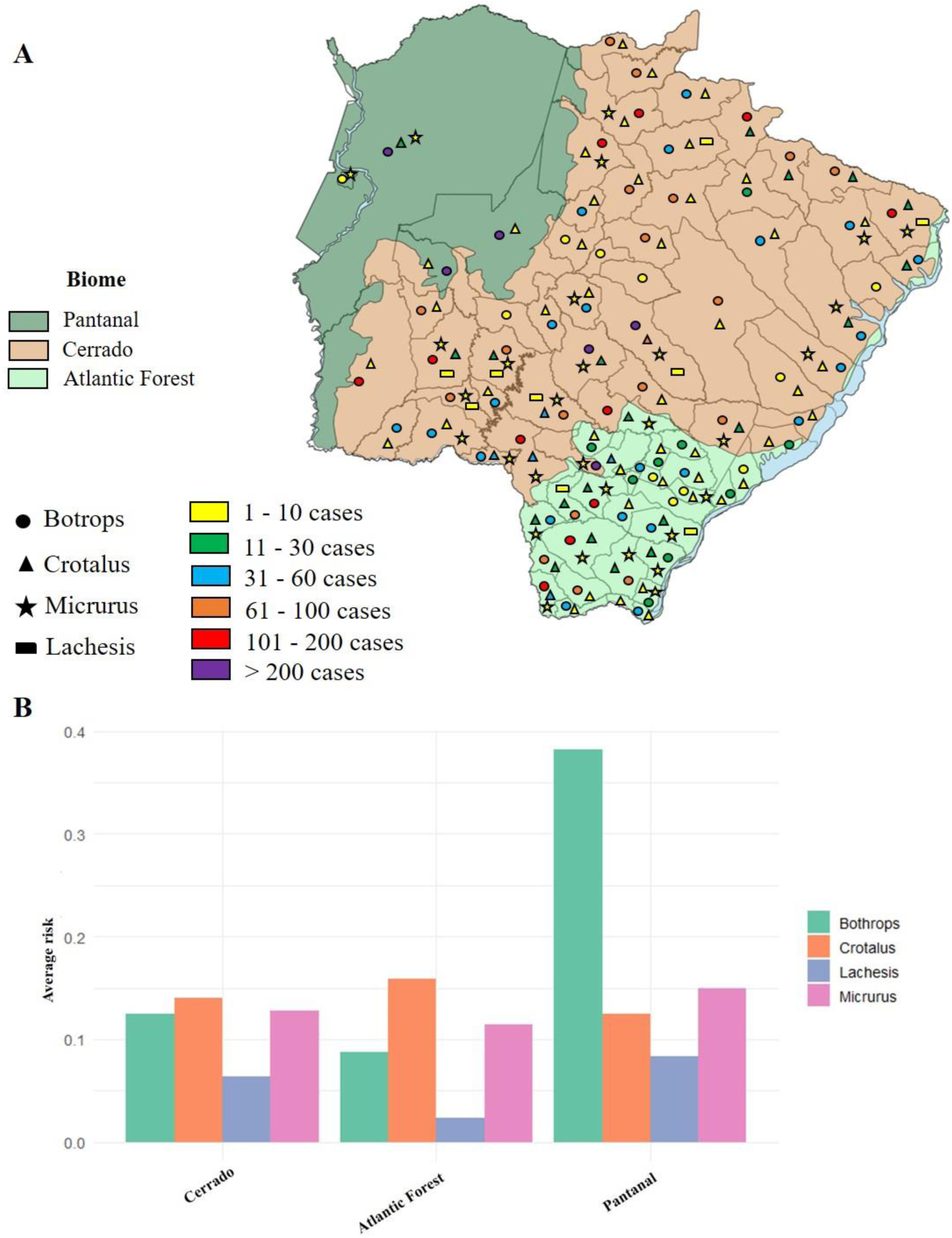
Snakebite envenomings by biome and snake genus in Mato Grosso do Sul (2007–2023). (A) Map of relative risk according to the predominant biome in each municipality, disaggregated by genus. Higher risk associated with *Bothrops* was observed in the Pantanal, while *Crotalus*, *Micrurus*, and *Lachesis* showed similar risks across the Cerrado and Atlantic Forest. *Lachesis* tended to present slightly lower risk in the Atlantic Forest. (B) Variation in total recorded cases among the Pantanal, Cerrado, and Atlantic Forest biomes by genus. No significant differences were detected (*Bothrops*: χ² = 5.15; df = 2; p = 0.076; *Crotalus*: χ² = 0.09; p = 0.958; *Micrurus*: χ² = 1.15; p = 0.564; *Lachesis*: χ² = 1.53; p = 0.467). Despite statistical non-significance, visual patterns suggest higher *Bothrops* occurrence in the Pantanal, possibly related to local ecological and environmental conditions.

**Figure S5.**
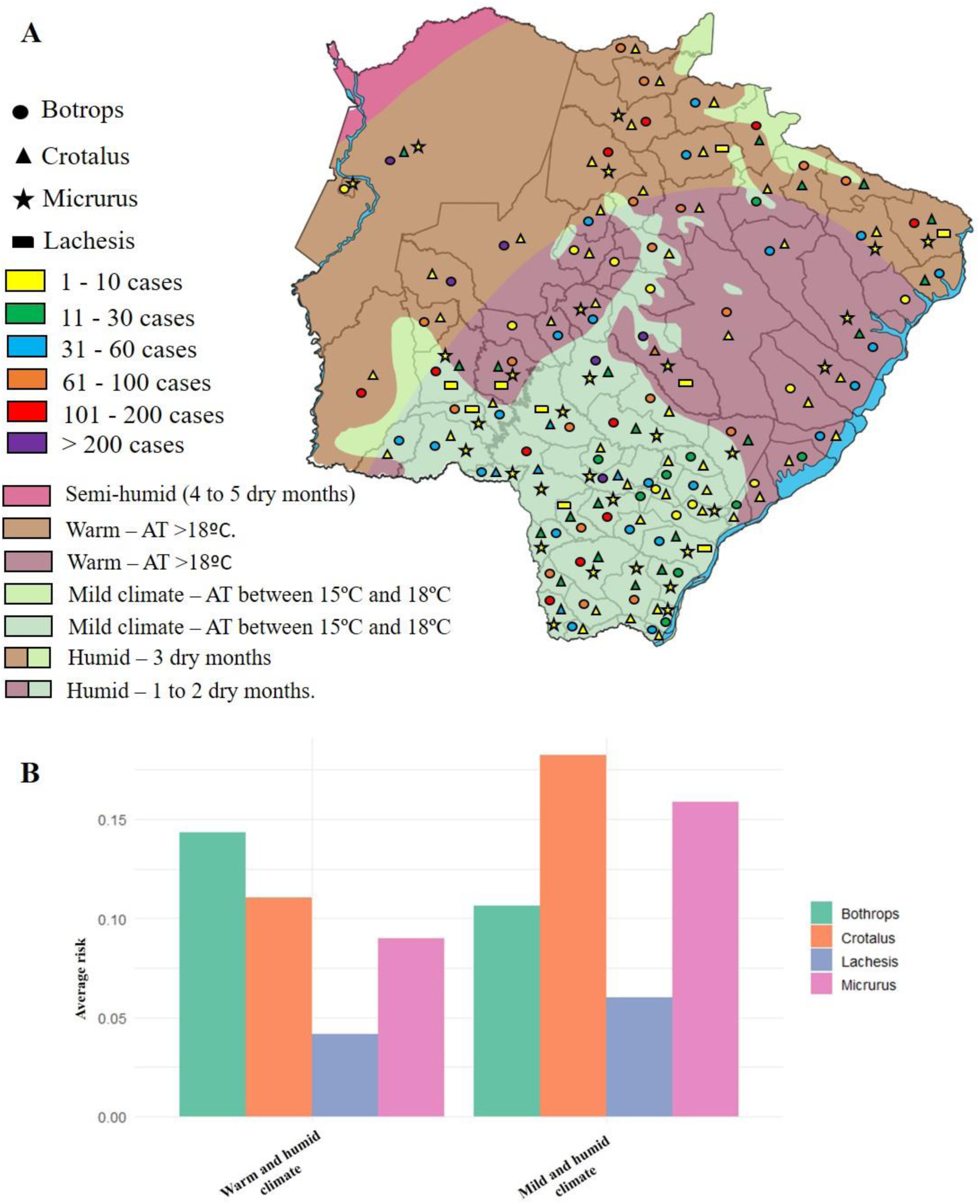
Snakebite envenomings by climatic type and snake genus in Mato Grosso do Sul (2007–2023). (A) Map of relative risk according to predominant climatic categories in the state. *Crotalus* and *Micrurus* showed relatively higher risk in hot semi-humid areas, whereas *Bothrops* and *Lachesis* displayed a more homogeneous distribution across climate types. (B) Variation in total cases among climatic types by genus. Kruskal–Wallis tests indicated no statistically significant differences (*Bothrops*: χ² = 0.30; p = 0.586; *Crotalus*: χ² = 1.85; p = 0.173; *Micrurus*: χ² = 1.68; p = 0.195; *Lachesis*: χ² = 0.01; p = 0.932). Although visual patterns suggest a possible climatic influence, especially for *Crotalus* and *Micrurus*, the differences were not statistically robust.

**Figure S6.**
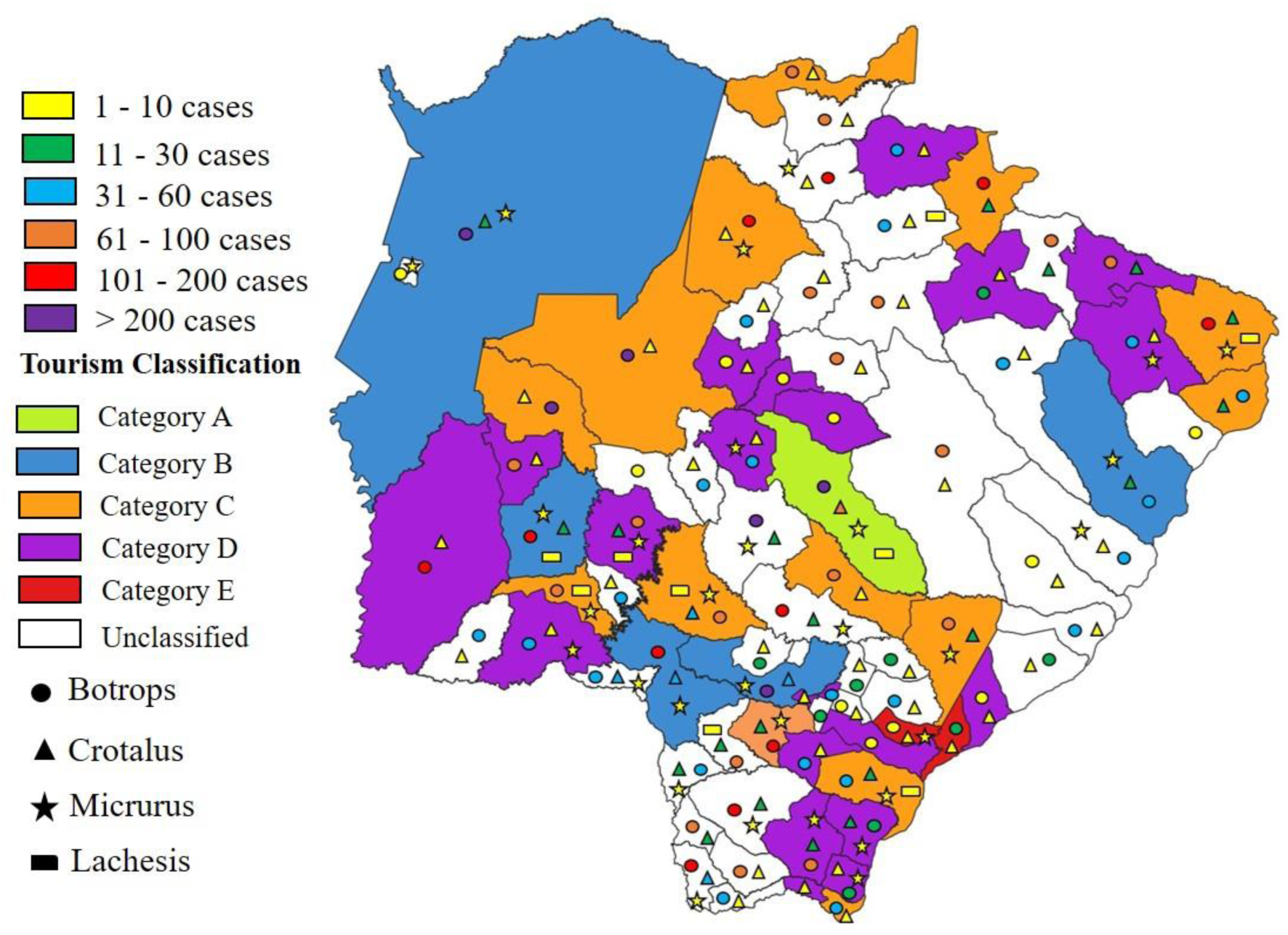
Spatial distribution of snakebite envenomings according to the tourism categorization of municipalities in Mato Grosso do Sul. Tourism-classified municipalities showed higher absolute numbers of cases, particularly in categories A and B. Relative risk was also higher for *Bothrops*, *Crotalus*, and *Micrurus* in these municipalities, suggesting potential influence of recreational and ecotourism activities on snakebite envenomings.

**Figure S7.**
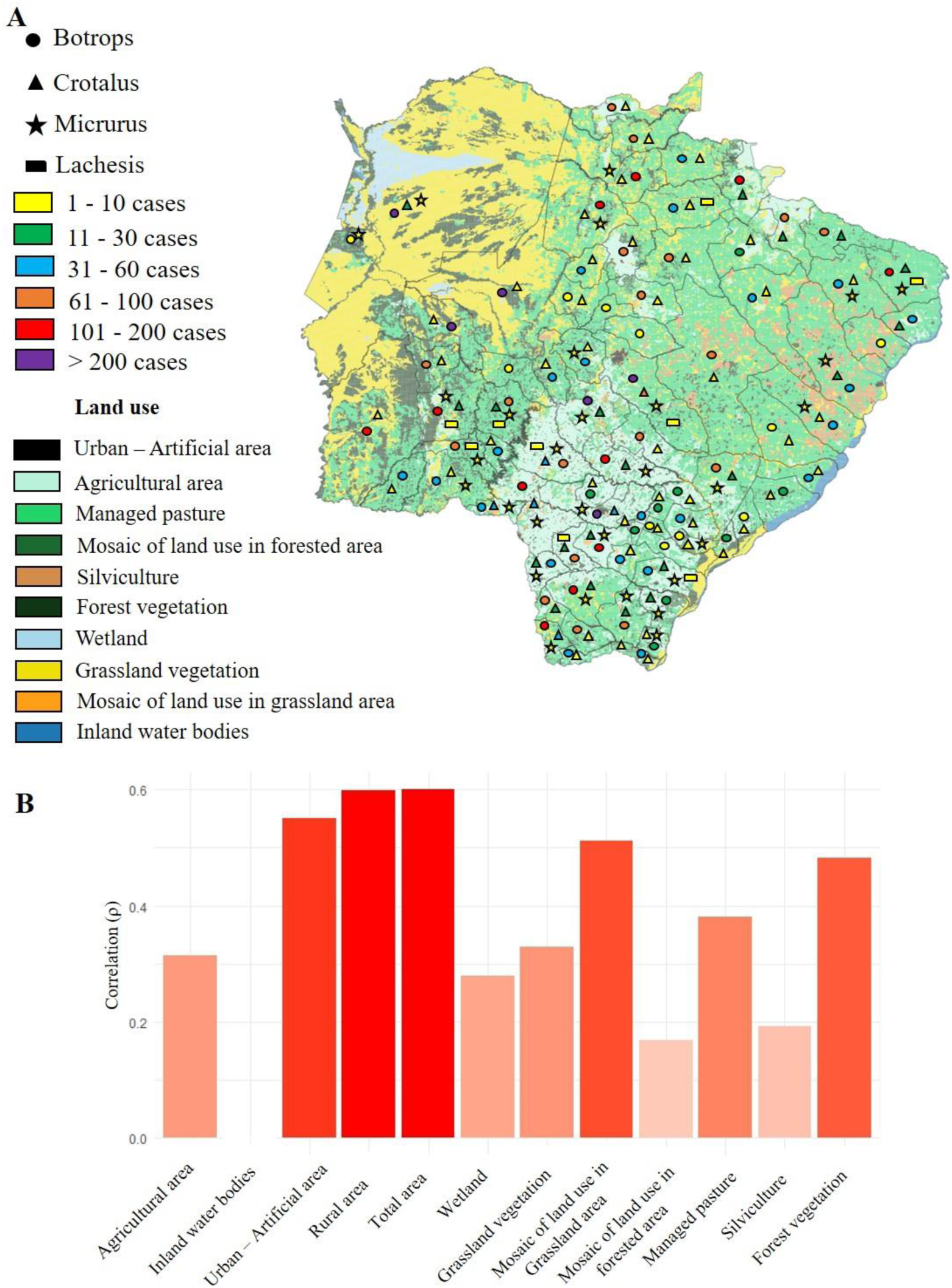
Relationship between land-use patterns and snakebite envenomings in Mato Grosso do Sul (2007–2023). (A) Distribution of snakebite cases according to major land-use and land-cover classes: artificial area, agriculture, pasture, forest mosaic, silviculture, forest vegetation, wetlands, grasslands, grassland mosaic, and continental water bodies. Higher case frequency was observed in agricultural and pasture areas, indicating direct influence of rural and livestock activities on human exposure. (B) Spearman’s correlation (ρ) between each land-cover type and total number of accidents. Total area, artificial area, grassland mosaic, and forest vegetation showed moderate-to-strong positive correlations, whereas agriculture, pasture, and grassland areas showed weaker positive associations. Continental water bodies were not significantly correlated. These findings reinforce the link between land-use patterns and snakebite distribution, suggesting that environmental changes and productive activities act as important determinants of risk.

**Table S1.** Multiple comparisons between different anatomical bite sites in snakebite envenomings in Mato Grosso do Sul (2007–2023). The table presents the results of Dunn’s multiple comparison test with Bonferroni correction, performed following the chi-square test, comparing the proportions of snakebite incidents across anatomical sites (head, trunk, arms, hands, legs, and feet). All pairwise comparisons were statistically significant (p < 0.001), except for a few residual combinations without relevant differences. These results highlight the predominance of bites on the lower limbs and hands, consistent with the typical occupational exposure observed in agricultural and rural settings.

| Body area -1 | Body area - 2 | p-value | CI - Low | CI – High |
| --- | --- | --- | --- | --- |
| Head | Arm | 1.074764e-65 | -0.5458 | -0.4443 |
| Head | Hand | 0.0 | -0.9084 | -0.8851 |
| Head | Torso | 1.127722e-19 | 0.3573 | 0.5379 |
| Head | Legs/feet | 0.0 | -0.9749 | -0.9684 |
| Head | Unclassified | 0.0 | -0.8415 | -0.8021 |
| Arm | Hand | 0.0 | -0.7395 | -0.7053 |
| Arm | Torso | 6.484927e-133 | 0.7306 | 0.8127 |
| Arm | Legs/feet | 0.0 | -0.9236 | -0.913 |
| Arm | Unclassified | 5.052028e-266 | -0.5772 | -0.5246 |
| Hand | Torso | 0.0 | 0.9516 | 0.967 |
| Hand | Legs/feet | 0.0 | -0.5917 | -0.572 |
| Hand | Unclassified | 3.699598e-155 | 0.2645 | 0.3052 |
| Torso | Legs/feet | 0.0 | -0.9912 | -0.987 |
| Torso | Unclassified | 0.0 | -0.9415 | -0.9146 |
| Legs/feet | Unclassified | 0.0 | 0.735 | 0.752 |

**Table S2.**
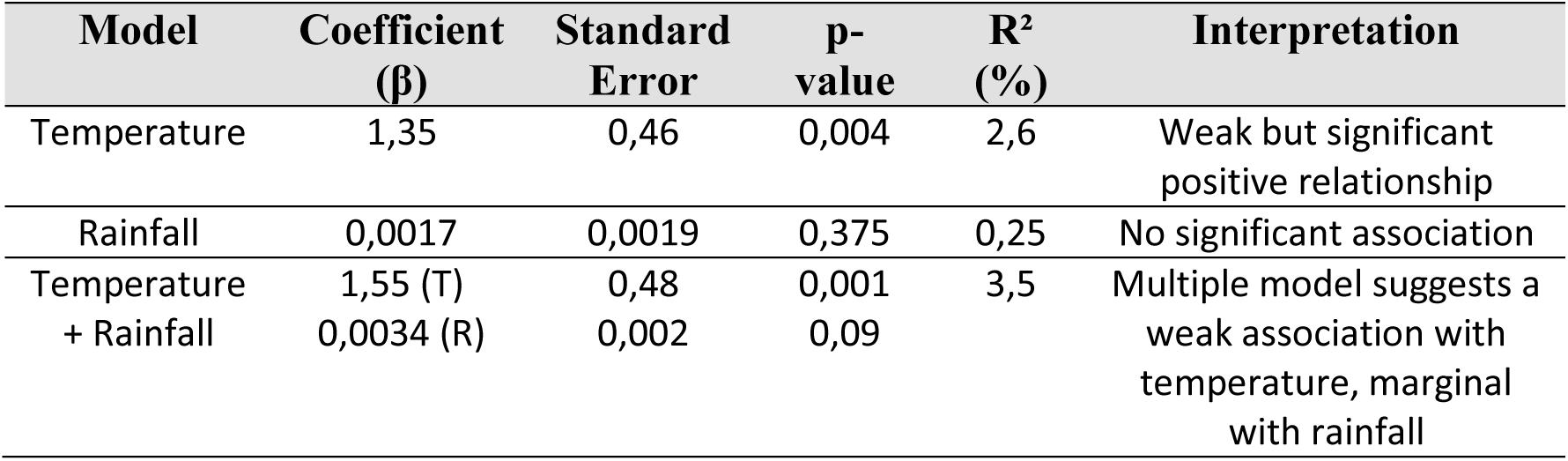
Association between climatic variables (mean annual temperature and precipitation) and the number of snakebite envenomings in Mato Grosso do Sul (2007–2023). Exploratory analyses indicated a positive, though weak, association between mean annual temperature and the number of recorded cases (β = 1.35; p = 0.004; R² = 2.6%), while precipitation showed no statistically significant relationship (p = 0.375). The multiple regression model retained the significance of temperature after adjustment for precipitation, but the overall explanatory power remained low (R² = 3.5%). These findings suggest a limited influence of climatic variables on snakebite occurrence, possibly reflecting the multifactorial complexity of environmental determinants.

| Model | Coefficient ($\beta$) | Standard Error | p-value | $R^2$ (%) | Interpretation |
| --- | --- | --- | --- | --- | --- |
| Temperature | 1,35 | 0,46 | 0,004 | 2,6 | Weak but significant positive relationship |
| Rainfall | 0,0017 | 0,0019 | 0,375 | 0,25 | No significant association |
| Temperature + Rainfall | 1,55 (T)<br>0,0034 (R) | 0,48<br>0,002 | 0,001<br>0,09 | 3,5 | Multiple model suggests a weak association with temperature, marginal with rainfall |

**Table S3.** Integrated model combining tourism and land-use variables to explain the spatial variation of snakebite cases in Mato Grosso do Sul. The model integrating tourism and land-use variables showed a fit comparable to the global model (AIC = 833.5 vs. 835.2), suggesting that biome and climate factors did not add significant explanatory power. Among land-use variables, agricultural areas (p = 0.007) and pastures (p = 0.029) remained positively associated with the number of cases, while artificial areas showed only a marginal trend (p = 0.084). Tourism categories exhibited high but non-significant coefficients when analyzed jointly with land-use factors, indicating overlapping effects. Overall, these results emphasize land occupation, particularly agriculture and pastures, and tourism flow as the main drivers influencing the spatial distribution of snakebite incidents across the state.

| Variable | Estimate (Global) | p-value (Global) | Estimate (Tourism + Soil) | P-value (Tourism + Soil) |
| --- | --- | --- | --- | --- |
| **Intercept** | -2.035 | 0.560 | -0.548 | 0.875 |
| **Tourism Category B** | 5.585 | 0.074 | 4.058 | 0.186 |
| **Tourism Category C** | 5.838 | 0.091 | 4.139 | 0.224 |
| **Tourism Category D** | 5.501 | 0.115 | 3.841 | 0.267 |
| **Tourism Category E** | 4.709 | 0.188 | 3.237 | 0.363 |
| **Tourism Category Unclassified** | 5.776 | 0.098 | 4.116 | 0.233 |
| **Biome: Atlantic Forest** | -0.079 | 0.755 | — | — |
| **Biome: Pantanal** | 0.073 | 0.852 | — | — |
| **Climate: Sub-humid warm** | -0.346 | 0.156 | — | — |
| **Artificial area (km<sup>2</sup>)** | 0.0298 | 0.024 * | 0.0226 | 0.084 |
| **Agricultural area (km<sup>2</sup>)** | 0.00020 | 0.160 | 0.00034 | 0.007 ** |
| **Pasture area (km<sup>2</sup>)** | 0.00016 | 0.026 * | 0.00015 | 0.029 * |
| **Mosaic in Forest area (km<sup>2</sup>)** | 0.00057 | 0.703 | -0.00017 | 0.909 |
| **Silviculture (km<sup>2</sup>)** | -0.00044 | 0.128 | -0.00046 | 0.114 |
| **Forest Vegetation (km<sup>2</sup>)** | 0.00015 | 0.294 | 0.00013 | 0.403 |
| **Wetland area (km<sup>2</sup>)** | -0.00102 | 0.158 | -0.00103 | 0.158 |
| **Grassland area (km<sup>2</sup>)** | 0.00012 | 0.297 | 0.00013 | 0.263 |
| **Mosaic in Grassland area (km<sup>2</sup>)** | -0.00038 | 0.707 | -0.00061 | 0.550 |
| **Inland Water** | -0.00135 | 0.246 | -0.00091 | 0.413 |
| **AIC** | 835.2 |  | 833.5 |  |

